# The tryptophan catabolite or kynurenine pathway in Alzheimer’s disease: a systematic review and meta-analysis

**DOI:** 10.1101/2022.03.18.22272608

**Authors:** Abbas F. Almulla, Thitiporn Supasitthumrong, Arisara Amrapala, Chavit Tunvirachaisakul, Al-Karrar Kais Abdul Jaleel, Gregory Oxenkrug, Hussein K. Al-Hakeim, Michael Maes

## Abstract

**Background:** Alzheimer’s disease (AD), characterized by gradual brain dysfunction and memory loss, is one of the major elderly health issues worldwide. Neuroinflammation and increased oxidative stress contribute to the pathophysiology of AD, thereby presumably inducing tryptophan (TRP) degradation through the TRP catabolite (TRYCAT) pathway.

**Objectives:** To delineate the activity of the TRYCAT pathway along with levels of TRP and tryptophan catabolites (TRYCATs) in AD patients.

**Methods:** We employed PubMed, Google Scholar, Web of Science, and SciFinder to obtain the relevant articles through a search process lasting the entire February 2022. We found 19 eligible articles which involved 738 patients and 665 healthy controls.

**Results:** Our result revealed a significant difference (*p* = 0.008) in the kynurenine (KYN)/TRP ratio (standardized mean difference, SMD = 0.216, 95% confidence interval, CI: 0.057; 0.376), and a significant decrease in TRP in AD patients (SMD = -0.520, 95% CI: -0.738; -0.302, p < 0.0001). Moreover, we also found a significant increase in the central nervous system (CNS, brain and cerebrospinal fluid, CSF) kynurenic acid (KA)/KYN ratio but not in peripheral blood, as well as a significant decrease in plasma KA and xanthurenic acid (XA) in the CNS and blood.

**Conclusions:** AD is characterized by TRP depletion but not by an overactivity of the TRYCAT pathway. IDO-induced production of neurotoxic TRYCATs is not a key factor in the pathophysiology of AD.

## Introduction

Alzheimer’s disease (AD) is one of the major elderly health issues worldwide, and is a progressive neurodegenerative disease with gradual memory deterioration and behavior aberrations (Reddy and Beal 2008, Cornutiu 2015, Morris, Berk et al. 2019). The pathophysiology of AD is associated with increased precipitation of amyloid-β (Aβ), aggregation of phosphorylated tau proteins, and apoptosis of neurons in the central nervous system (CNS) (Mattson 2004, LaFerla, Green et al. 2007, O’Brien and Wong 2011). However, these pathophysiological findings are possibly non-unique contributors to the AD pathophysiology because normal individuals without cognitive symptoms may also exhibit elevations in amyloid deposits, as indicated by positron emission tomography studies (Chételat, La Joie et al. 2013). Moreover, clinical severity in AD patients is not significantly correlated with amyloid deposition. Hence, the pathophysiological picture of AD remains ambiguous (Chételat, La Joie et al. 2013, Morris, Clark et al. 2014).

Several postmortem studies have indicated the implication of an imbalanced immune system in the pathological state of AD. These studies reported that amyloid plaques in the brain tissues of AD patients showed an aggregate of interleukin (IL)-1β (Griffin, Stanley et al. 1989), IL-6 (Hüll, Berger et al. 1996, Bonaccorso, Lin et al. 1998), and tumor growth factor-β (TGF-β) (van der Wal, Gómez-Pinilla et al. 1993). Thus, AD is associated with neuroinflammation followed by upregulated microglia and astrocytes, and increased neuroinflammatory markers (Heneka, Carson et al. 2015). In addition, in cerebrospinal fluid (CSF) and plasma of AD patients, elevated levels of pro and anti-inflammatory cytokines are observed, namely IL-1β, IL-6, and tumor necrosis factor (TNF)-α, and IL-1 receptor antagonist (IL-1ra) and IL-10, respectively (Maes, DeVos et al. 1999, Brosseron, Krauthausen et al. 2014). High plasma levels of cortisol and interferon-gamma (IFN-γ) were found especially in the milder phases of AD patients (Zvěřová, Fišar et al. 2013, Belkhelfa, Rafa et al. 2014).

Neuronal cells are strongly influenced by oxidative and nitrosative stress (O&NS), much more than peripheral tissues, due to the increased rate of metabolism and relative lower levels of antioxidants (Maes, Galecki et al. 2011). Moreover, amyloid plaque aggregations cause neurotoxicity because they drive microglia and astrocytes overactivity, releasing pro-inflammatory cytokines and reactive oxygen species (ROS), consequently causing O&NS in the brain (Moneim 2015). Administration of antioxidants may help prevent disease progression and decrease risk of developing AD (Moneim 2015). Furthermore, in AD patients, a large body of evidence has confirmed the presence of increased plasma levels of lipopolysaccharides (LPS) (Zhang, Miller et al. 2009, Zhan, Stamova et al. 2016, Zhao, Jaber et al. 2017, Kim, Kim et al. 2021), which intrinsically has toxic effects and promotes O&NS and immune activation (Halawa, A El-Adl et al. 2018).

Immune activation with increased production of IFN-γ and M1 macrophage cytokines, and increased ROS/O&NS and LPS may induce indoleamine 2,3-dioxygenase, the key regulatory enzyme in the tryptophan catabolite (TRYCAT) pathway (Maes, Leonard et al. 2011). IDO activation may cause a reduction in tryptophan (TRP) levels and increased TRYCAT levels as shown in **Figure 1** (Maes, Meltzer et al. 1993, Smith and Maes 1995, Kanchanatawan, Sirivichayakul et al. 2018). Indeed, studies on AD and mild cognitive impairment (MCI) patients display a high kynurenine (KYN)/TRP ratio (Widner, Leblhuber et al. 2000, Greilberger, Fuchs et al. 2010). Thus, TRP will be diverted from the production of vital derivates, such as serotonin, which maintains neuroplasticity and protection against neuronal damage (Croonenberghs, Verkerk et al. 2005, Radulescu, Dragoi et al. 2021) to the TRYCAT pathway, potentially producing neurotoxic metabolites, including quinolinic acid (QA), xanthurenic acid (XA), 3-hydroxykynurenine (3HK), picolinic acid (PA), and 3-hydroxyanthranilic acid (3HA).

**Figure 1:**
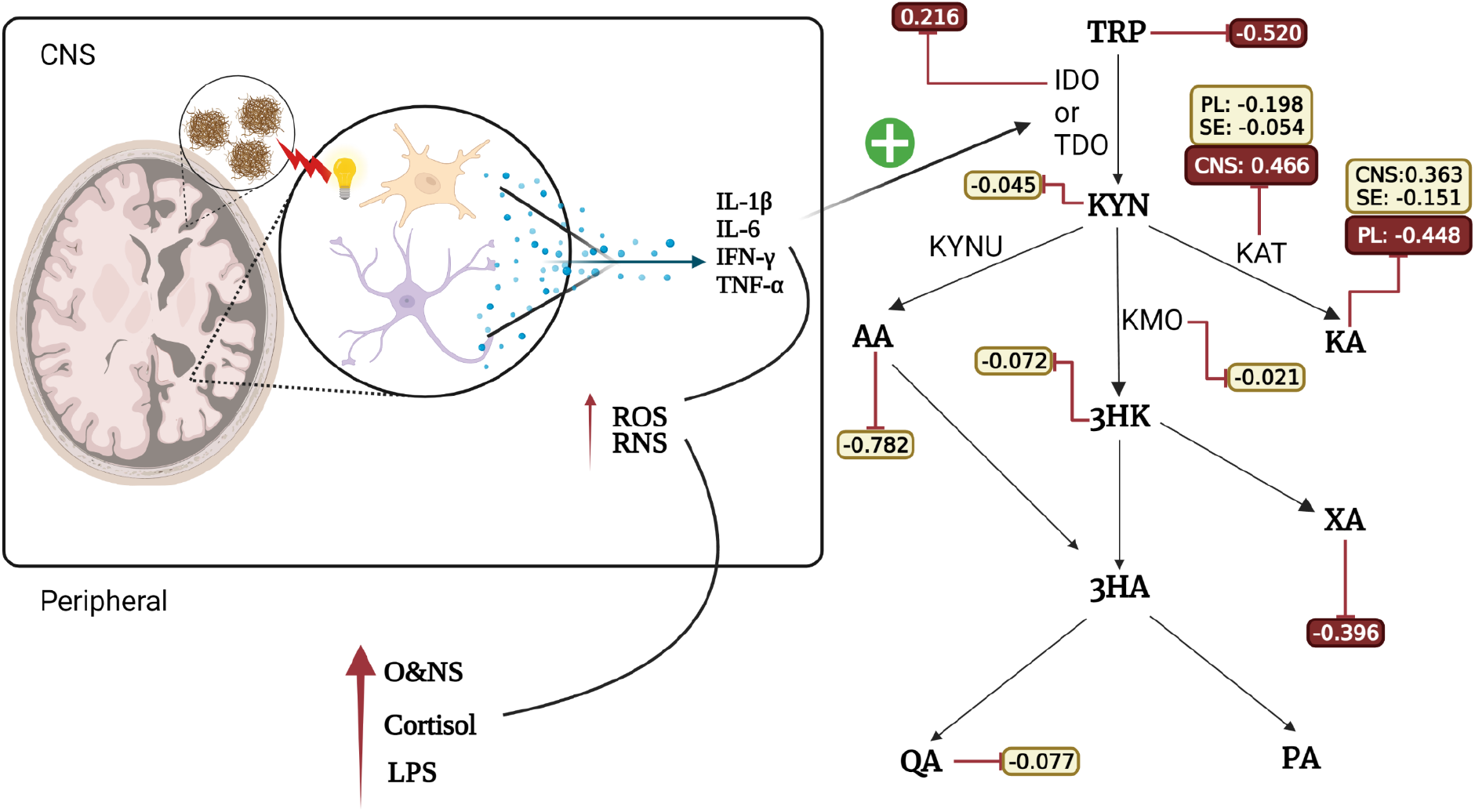
Summary of the TRYCAT pathway in AD IFN-γ: Interferon-Gamma, IL-6: Interleukin 6, IL-1β: Interleukin-1 beta, O&NS: Oxidative and nitrosative stress, LPS: Lipopolysaccharides, CNS: Central nervous system, IDO: Indoleamine 2,3 dioxygenase, TDO: Tryptophan 2,3-dioxygenase, KAT: Kynurenine Aminotransferase, KMO: Kynurenine 3-monooxygenase, KYNU: Kynureninase, TRP: Tryptophan, KYN: Kynurenine, KA: Kynurenic Acid, 3HK: 3-Hydroxykynurenine, AA: Anthranilic Acid, XA: Xanthurenic Acid, 3HA: 3-Hydroxyanthranilic Acid, PA: Picolinic Acid, QA: Quinolinic Acid.

Some of these tryptophan catabolites (TRYCATs), namely 3HA, 3HK, and QA, are pro-oxidants and induce O&NS via generation of ROS and reactive nitrogen species (RNS) such as superoxide ion and peroxides, whilst also boosting copper-dependent alpha-crystallin cross-linking (Goldstein, Leopold et al. 2000, Smith, Smith et al. 2009, Zadori, Veres et al. 2018). Moreover, these metabolites mediate cell damage due to the formation of free radicals (Dykens, Sullivan et al. 1987, Okuda, Nishiyama et al. 1998). QA may also cause lipid peroxidation as a result of producing ROS (Rios and Santamaria 1991, Santamaría, Galván-Arzate et al. 2001) and has been reported to exhibit excitatory properties on N-methyl-D-aspartate (NMDA) receptors (Kanchanatawan, Sirivichayakul et al. 2018) and pro-inflammatory effects, particularly in the microglia thereby augmenting the inflammatory, oxidative stress and neurotoxicity effects (Maes, Leonard et al. 2011).

On the other hand, various TRYCATs also have neuroprotective roles, such as kynurenic acid (KA), which regulates NMDA receptor functions (Morris, Carvalho et al. 2016), whilst anthranilic acid (AA) impedes the production of QA and PA (Guillemin, Cullen et al. 2007, Almulla and Maes 2022). Furthermore, XA may have neuroprotective effects via reducing vesicular glutamate transport (VGLUT), downregulating NMDA receptors, and attenuating postsynaptic excitatory potency (Kanchanatawan, Sirivichayakul et al. 2018).

Nonetheless, in AD there is no systematic review and meta-analysis examining TRYCAT pathway activity. Hence, in the current study, we perform a comprehensive meta-analysis and systematic review of TRP and TRYCAT data in patients with AD compared to healthy controls.

## Material and Methods

We employed the Preferred Reporting Items for Systematic Reviews and Meta-Analyses (PRISMA) 2020 criteria, the Cochrane Handbook for Systematic Reviews and Interventions guidelines, and Meta-Analyses of Observational Studies in Epidemiology (MOOSE) guidelines to conduct the present meta-analysis. We investigate TRP along with TRYCATs levels, whilst IDO, KAT, and KMO enzyme activity were examined through their indices, namely, KYN/TRP and (KA+KYN)/TRP, KA/KYN and KA/(KYN+TRP), and 3HK/KYN ratios, respectively.

### Search strategy

We began the database search process on January 1^st^, 2022. Specific keywords and AD-associated MESH terms were entered into the electronic databases, namely PubMed/MEDLINE, Google Scholar, Web of Science, and SciFinder. These keywords, along with the terms, covered TRP, TRYCATs, and AD, all of which are displayed in the electronic supplementary file (ESF), Table 1. The reference lists of prior reviews and grey literature were also examined to ensure that all relevant papers were included. By the end of the month, all relevant articles were collected.

**Table 1.**
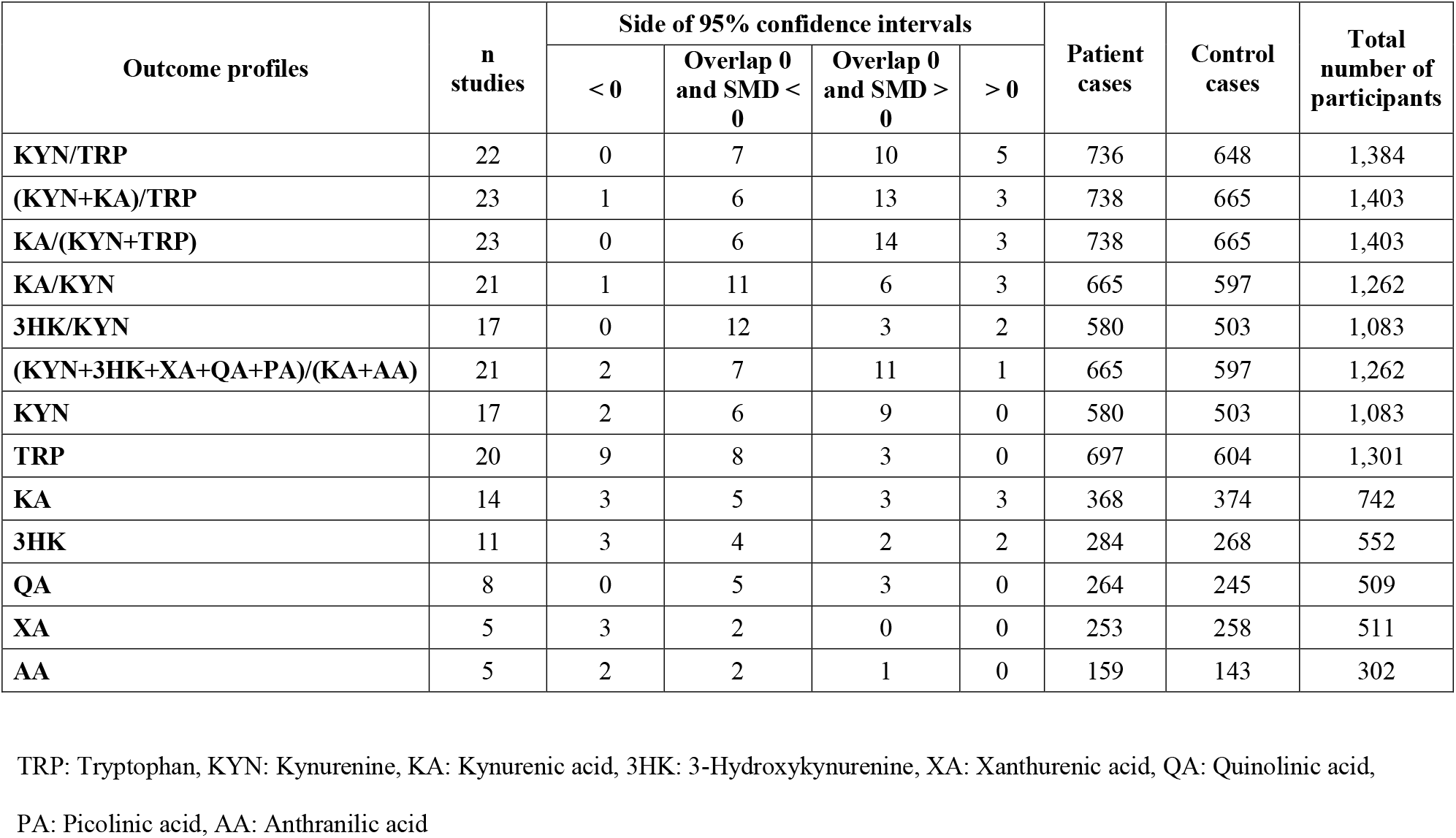
The outcomes profile and number of AD patients and healthy control along with the side of standardized mean difference (SMD) and the 95% confidence intervals with respect to zero SMD

### Eligibility Criteria

Peer-reviewed journals and English language were the primary criteria for selecting eligible articles for the study. We did, nevertheless, assess manuscripts in other languages, including Thai, French, Spanish, German, Italian, and Arabic. All included articles were compliant with the following inclusion criteria: a) observational case-control studies that employed serum, plasma, CSF, and brain tissues to investigate the levels of TRP and TRYCATs in patients with AD compared to healthy controls; b) studies that recruited cognitively normal individuals as a control group and who were free of any active psychiatric illness; and c) articles that presented numerical values of mean with standard deviation (SD) or mean standard error (SEM) for TRP and TRYCATs. Based on the specified criteria, the following studies were excluded a) systematic and narrative review articles, as well as meta-analyses; b) animal and genetic studies, and duplicate articles; c) studies which used saliva and hair to assay TRYCATs; d) studies reporting other types of dementia or pre-clinical AD patients; and e) articles that did not reveal mean SD/SE of the assessed TRP and TRYCATs. We emailed the authors to request the mean and SD of the biomarkers if they published geometric means, median (interquartile range, range), or showed data as a graph. In the absence of a response from the authors, we used Wan and Wang’s estimation method to compute mean SD from the median and interquartile range or range. Furthermore, Web Plot Digitizer was used to extract quantitative data from graphs showing means and SD/SEM of TRYCATs (https://automeris.io/WebPlotDigitizer/).

### Primary and secondary outcomes

The KYN/TRP and (KYN+KA)/TRP ratios were utilized to examine the primary outcome, which was IDO enzyme activity in AD patients versus healthy controls. Secondary outcomes included the KAT enzyme indices, namely KA/KYN and KA/(KYN+TRP) ratios, the KMO enzyme proxy 3HK/KYN ratio, and the neurotoxic index (KYN+3HK+QA+XA+PA)/(KA+AA). The levels of TRP, KYN, KA, 3HK, AA, XA, and QA were also assessed in patients with AD versus healthy controls.

### Screening and data extraction

The first author (AA) carried out the preliminary review to determine eligible studies for inclusion by inspecting the title and abstract of the searched articles. The full texts of the eligible studies were then downloaded after screening with the pre-specified exclusion criteria. After the first author finished inputting the data in an Excel spreadsheet, the second author (TS) reviewed the accuracy of the data. The last author (MM) was consulted for any equivocal findings. A pre-specified excel spreadsheet was used to extract and organize the required data in this study. This data file comprised the authors’ names, publication dates, numerical data of TRP and TRYCATs, the number of recruited individuals either as AD patients or healthy controls, background variables such as age and number of males and females in each group, statistical data such as mean (SD/SE), sample type, mini-mental state examination (MMSE) score, the latitude of the country where the study was conducted, and the quality and redpoint scores of the included studies. The methodological quality of included studies was evaluated using the prior known “immune confounder scales (ICS)” after certain modifications were made by the last author (MM), as shown in ESF, Table 3. We assessed the essential quality determinants by the ICS, namely sample size, controlling of the confounders, and the time at which the sample was taken. The range of the overall quality score was 0 to 10, with 10 being the best quality. An uncontrolled study design and insufficient adjustment of major confounders were addressed in the redpoint score scale, which has a range of 0 to 26, with higher scores implicating a lack of control and higher exposure of the TRYCATs results to many types of bias, including biological and analytical.

### Data analysis

We performed the present meta-analysis using the CMA V3 software and complied with all PRISMA criteria. As a minimum, three studies reporting each TRYCAT was required to carry out a meta-analysis. **Table 1** shows the outcome profile of the TRYCATs, which were addressed in the current systematic review and meta-analysis. When several biomarkers representing the same profile were reported in the same research, a synthetic score was calculated using the mean of the results, assuming dependency. The computed scores, which reflect different profiles, as well as solitary TRYCATs were compared between AD patients and healthy controls. We examined the activity of IDO, KAT and KMO enzymes through their proxies as follows: a) IDO was assessed using KYN/TRP and (KYN+KA)/TRP ratios by setting the effect size direction of KYN (KA) as positive for increased KYN (KA) and positive for lowered TRP levels in AD; and b) KAT activity was determined by the KA/KYN and KA/(KYN+TRP) ratios by specifying a positive effect size direction for increased KA in AD patients and a negative direction for TRP and KYN in the meta-analysis. The random-effects model with restricted maximum-likelihood was utilized in this investigation on the premise that population characteristics varied between studies. The effect size was calculated using the standardized mean difference (SMD) with 95% confidence intervals (95% CI), and two-tailed, *p* < 0.05 was considered statistically significant. SMD values of 0.8, 0.5 and 0.2 indicate large, moderate, and modest effect sizes, respectively (Cohen 2013).

We computed tau-squared, as previously indicated, to evaluate the heterogeneity in addition to the Q and I^2^ values (Vasupanrajit, Jirakran et al. 2021, Vasupanrajit, Jirakran et al. 2021). To test the strength of the findings, we performed a sensitivity analysis by using the “removing-one-study approach.” Subgroup analysis was employed in this meta-analysis to examine the differences between brain tissues, CSF, serum, and plasma (Almulla, Vasupanrajit et al. 2021), and each of these groups were considered as the unit of analysis. In the case that there were no significant differences between brain tissues and CSF, they were combined to a single entity called the central nervous system (CNS). The fail-safe N method, Kendall tau with continuity correction, and Egger’s regression intercept (both using a one-tailed *p*-value) were used to reveal the influence of small study effects. We accounted for the modified effect size computed by imputing the missing studies by the Duval and Tweedie’s trim-and-fill method in case of significant asymmetry reported by Egger’s test. We conducted a meta-regression in the current study to assess the sources of heterogeneity.

## Results

### Search findings

PRISMA flowchart **Figure 2** shows the detailed results of the search processes in the different databases and the standards used for inclusion-exclusion of research articles. Based on prespecified terms and keywords listed in ESF, Table 1, we initially identified 24 full-text papers. Five articles were removed due to reasons mentioned in the ESF, Table 5. Accordingly, 19 full-text articles were considered in the current meta-analysis (Tohgi, Abe et al. 1992, Tohgi, Abe et al. 1995, Bonaccorso, Lin et al. 1998, Baran, Jellinger et al. 1999, Widner, Leblhuber et al. 2000, Hartai, Juhasz et al. 2007, Greilberger, Fuchs et al. 2010, Gulaj, Pawlak et al. 2010, Schwarz, Guillemin et al. 2013, Wennstrom, Nielsen et al. 2014, Rommer, Fuchs et al. 2016, Giil, Midttun et al. 2017, Oxenkrug, van der Hart et al. 2017, Jacobs, Lim et al. 2019, Sorgdrager, Vermeiren et al. 2019, van der Velpen, Teav et al. 2019, Gonzalez-Sanchez, Jimenez et al. 2020, Whiley, Chappell et al. 2021, Willette, Pappas et al. 2021).

**Figure 2:**
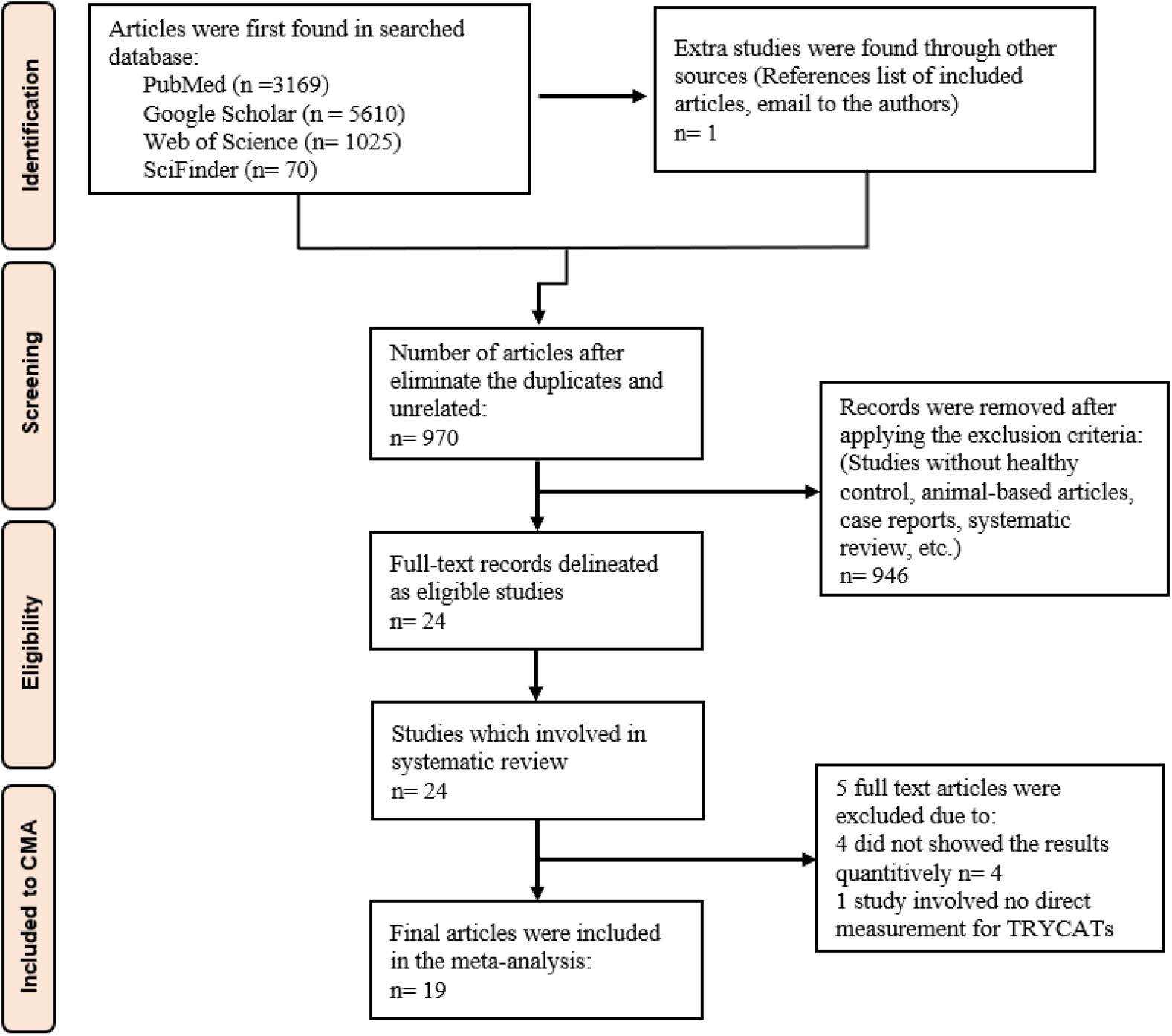
The PRISMA flow chart

We included case-control studies in this meta-analysis, resulting in a total investigation of 1,403 individuals, distributed as 738 AD patients and 665 healthy controls. We examined the subjects’ TRP and TRYCATs levels in 8 studies within the CNS (7 CSF and 1 brain tissue), 8 in plasma, and 7 in serum. Four studies measured central and peripheral levels of the TRP and TRYCATs. In the serum, TRP and TRYCAT levels were examined in 582 participants (321 AD patients versus 261 healthy controls), 487 participants (244 AD patients versus 243 healthy controls) were evaluated in plasma studies, and 335 participants (173 AD patients versus 162 healthy controls) were investigated in CNS studies. No significant differences were found between central and peripheral outcomes except in KA levels and KA/KYN ratio. Additionally, there were no significant differences between serum and plasma. Therefore, we combined all serum/plasma studies and pooled the overall effect size. High-performance liquid chromatography (HPLC) was the most used analytic technique (reported in 10 studies), followed by LC (2 studies), while the remaining studies employed different techniques, as listed in ESF, Table 5. Austria contributed with 4 studies, while Belgium, Japan, Sweden, and the USA participated with 2 studies, and 1 study was reported by research groups from Germany, Hungary, Norway, Poland, Switzerland, UK, and Spain. The redpoint and quality scores were 11 (median) (min=10, max=12.5) and 5.5 (min=2.5 max=9), respectively. The subjects’ ages ranged from 63 to 81 years.

### Primary outcome variables

#### KYN/TRP and (KYN+KA)/TRP ratios

In the current meta-analysis, we included 22 studies concerning the KYN/TRP ratio: 5 studies with 95% CI that were entirely on the positive side of zero, whereas no 95% CI were entirely on the negative side of zero. The 95% CI of the other 17 studies intersected with zero, with 10 studies showing a positive SMD value, and 7 studies showing SMD values less than zero as displayed in **Table 1**. The overall results in **Table 2** indicate a significant difference with modest effect size in the KYN/TRP ratio between AD patients and healthy controls, as illustrated in the forest plot (**Figure 3)**. Publication bias was observed in 2 studies to the left, and the adjusted estimate point changed to 0.165 (95%CI:-0.001;0.332) after imputing those 2 studies.

**Table 2.**
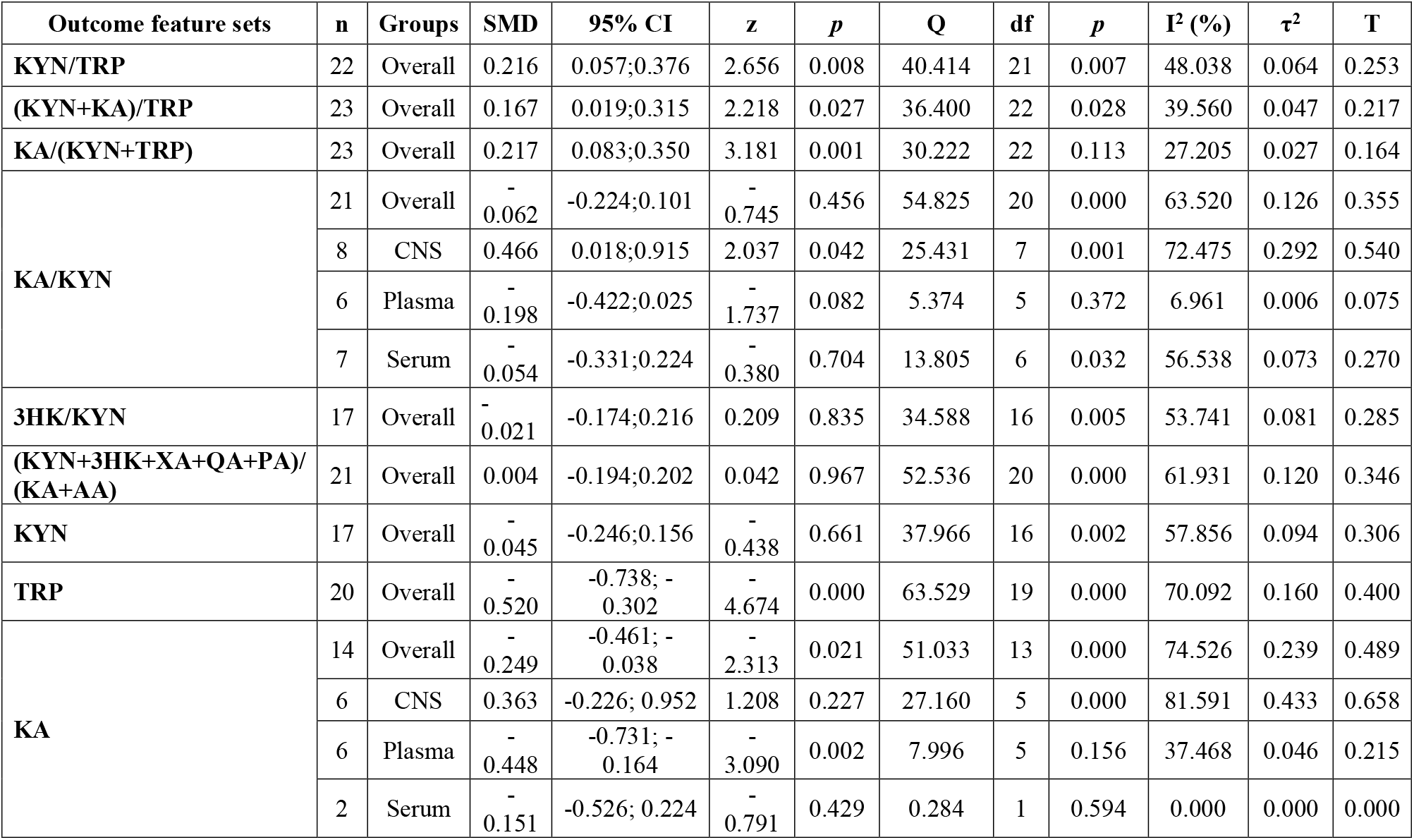
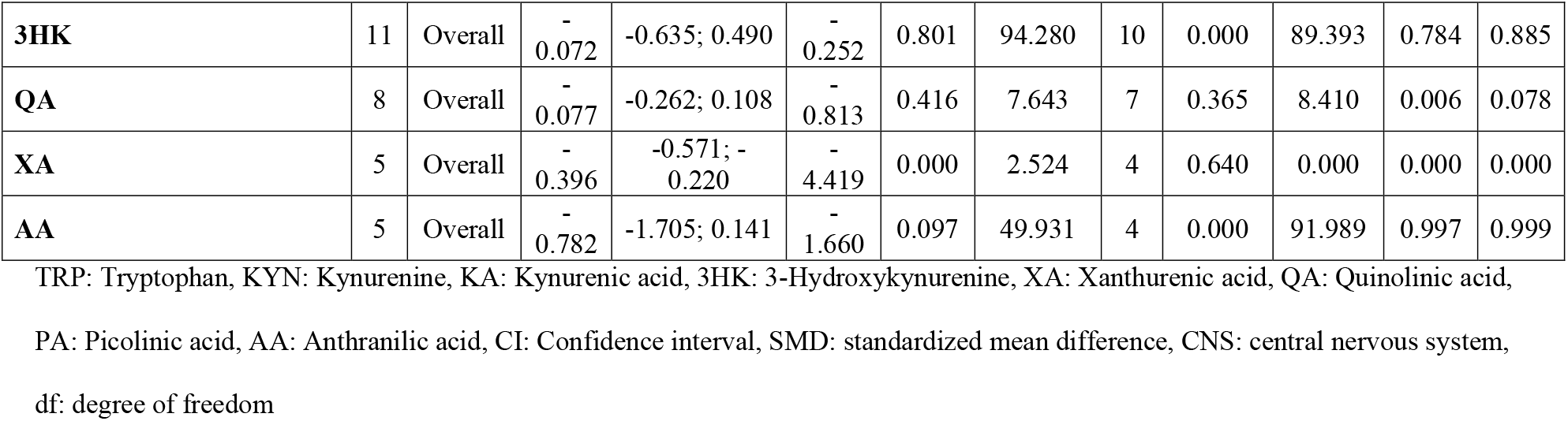
Results of meta-analysis performed on several outcomes (TRYCATs) variables with combined different media and separately.

**Figure 3:**
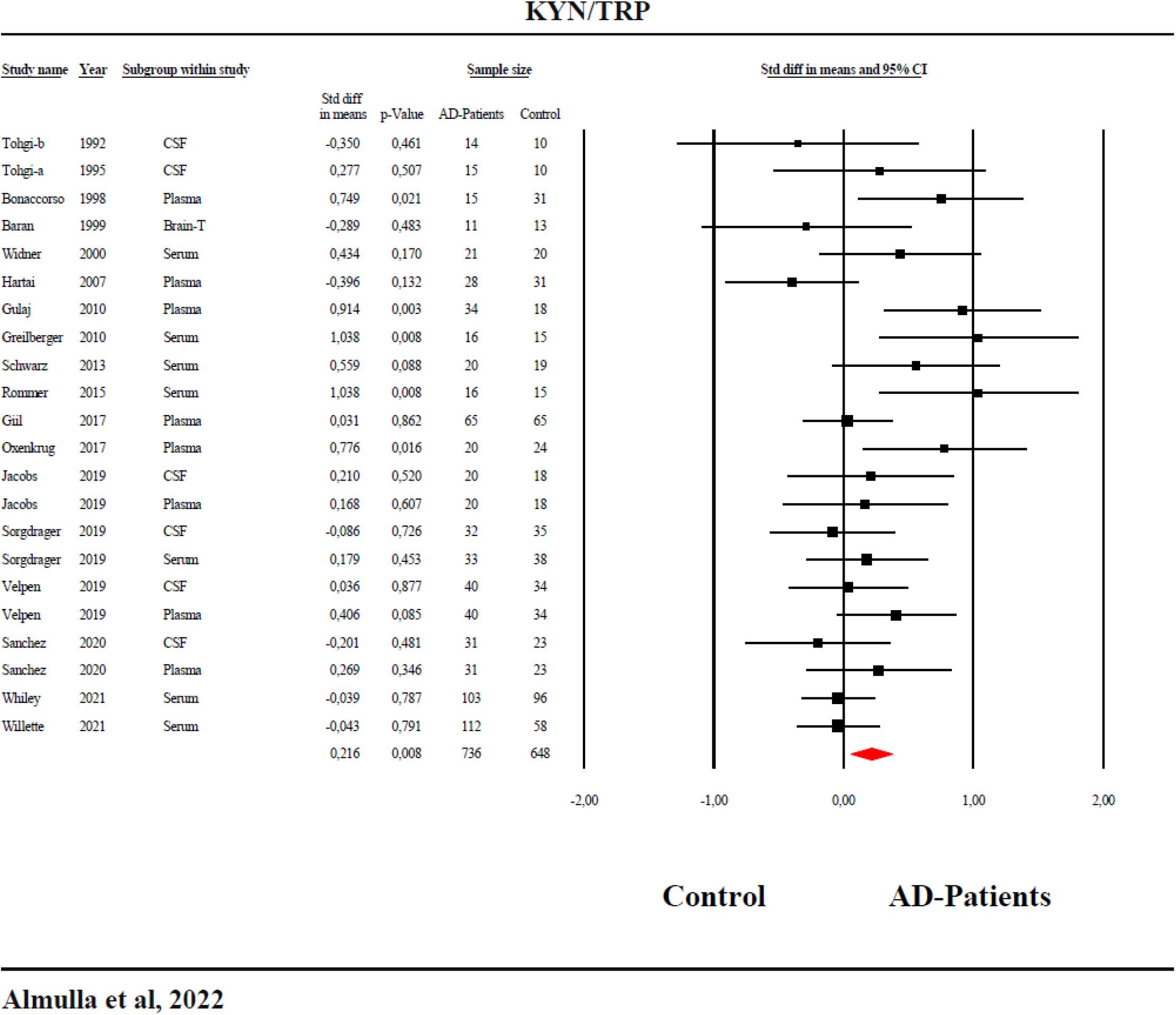
Forest plot of the kynurenine/tryptophan (KYN/TRP) ratio reflecting indoleamine-2,3-dioxygenase (IDO) activity with results of a meta-analysis conducted on 22 studies.

The forest plot for the (KYN+KA)/TRP ratio is shown in ESF, Figure 1. **Table 2** indicates a significant increase with a very modest effect size in AD compared to healthy controls. **Table 3** indicates a possible bias with 8 studies to the left side. Imputing these studies alters the SMD to 0.015 (95%CI: -0.177;0.147), which is no longer significant.

**Table 3.** Results on publication bias.

| **Outcome feature sets** | **Fail safe n** | **Z Kendall's $\tau$** | ***p*** | **Egger's t test (df)** | ***p*** | **Missing studies (side)** | **After adjustment** |
| --- | --- | --- | --- | --- | --- | --- | --- |
| **KYN/TRP (Overall)** | 3.651 | 1.889 | 0.029 | 2.353 (20) | 0.014 | 2 (Left) | SMD= 0.165;<br>95% CI (-0.001;0.332) |
| **(KYN+KA)/TRP (Overall)** | 2.985 | 2.033 | 0.021 | 2.672 (21) | 0.007 | 8 (Left) | SMD= 0.015;<br>95% CI (-0.177;0.147) |
| **KA/(KYN+TRP) (Overall)** | 4.095 | 2.561 | 0.005 | 1.532 (21) | 0.070 | 4 (Left) | SMD= 0.147;<br>95% CI (-0.000;0.294) |
| **KA/KYN (Overall)** | 0.923 | 0.966 | 0.166 | 0.002 (19) | 0.498 | 3 (Right) | SMD= 0.137;<br>95% CI (-0.063;0.338) |
| **KA/KYN (CNS)** | 3.521 | 1.608 | 0.053 | 2.513 (6) | 0.022 | 1 (Right) | SMD=0.615;<br>95% CI (0.112;1.118) |
| **KA/KYN (Plasma)** | -1.925 | 0.000 | 0.500 | 0.775 (4) | 0.240 | 1 (Left) | SMD= -0.224;<br>95% CI(-0.433; -0.015) |
| **KA/KYN (Serum)** | -0.381 | 2.252 | 0.012 | 3.818 (5) | 0.006 | 3 (Right) | SMD= 0.095;<br>95% CI(-0.149;0.339) |
| **3HK/KYN** | 0.405 | 1.071 | 0.142 | 1.119 (15) | 0.140 | 3 (Right) | SMD= 0.092;<br>95% CI(-0.094;0.280) |
| **(KYN+3HK+XA+QA+PA)/**<br>**(KA+AA)** | -0.022 | 0.241 | 0.404 | 0.689 (19) | 0.249 | 4 (Left) | SMD= -0.128;<br>95% CI(-0.334;0.078) |
| **KYN** | -0.981 | 0.576 | 0.282 | 0.977 (15) | 0.171 | 4 (Left) | SMD= -0.170;<br>95% CI(-0.368; 0.026) |
| **TRP** | -7.982 | 2.984 | 0.001 | 3.175 (18) | 0.002 | 2 (Right) | SMD= -0.432;<br>95% CI(-0.660; -0.204) |
| **KA (Overall)** | -1.274 | 1.970 | 0.024 | 1.832 (12) | 0.045 | 0 |  |
| **KA (CNS)** | 2.360 | 1.127 | 0.129 | 1.664 (4) | 0.085 | 0 |  |
| **KA (Plasma)** | -3.899 | 0.751 | 0.226 | 0.126 (4) | 0.452 | 0 |  |
| **3HK** | -0.924 | 0.311 | 0.377 | 0.441 (9) | 0.334 | 3 (Right) | SMD= 0.350;<br>95% CI(-0.245; 0.947) |
| **QA** | -0.673 | 0.123 | 0.450 | 1.006 (6) | 0.176 | 0 |  |
| **XA** | -4.513 | 1.224 | 0.110 | 1.306 (3) | 0.141 | 2 (Right) | SMD= -0.336;<br>95%CI (-0.495; -0.177) |
| **AA** | -4.584 | 0.244 | 0.403 | 0.974 (3) | 0.200 | 1 (Left) | SMD= -1.047;<br>95% CI(-1.993; -0.101) |

TRP: Tryptophan, KYN: Kynurenine, KA: Kynurenic acid, 3HK: 3-Hydroxykynurenine, XA: Xanthurenic acid, QA: Quinolinic acid, PA: Picolinic acid, AA: Anthranilic acid, CI: Confidence interval, SMD: standardized mean difference, CNS: central nervous system, df: degree of freedom

### Secondary outcome variables

#### The KA/KYN and KA/(KYN+TRP) ratios

We found a high heterogeneity in the KA/KYN results and, therefore, performed subgroup analyses that revealed a significant difference (*p*=0.034) between CNS and peripheral levels. **Table 2** and ESF, Figure 2 shows that the KA/KYN ratio in the CNS of AD patients significantly increased compared to healthy controls with a moderate effect size (SMD=0.466). Contrastingly, the results in serum and plasma were not significant. **Table 3** indicates a possible bias in the CNS results with 1 study to the right side missing and after imputation, the SMD increased to 0.615 (95%CI: 0.112; 1.118). The results in plasma and serum showed a possible bias with 1 study missing on the left side for plasma, and 3 studies on the right side for serum. After adjusting the effect sizes by imputing for those studies, the results remained significant.

The forest plot of the KA/(KYN+TRP) ratio (ESF, Figure 3) and **Table 2** show a significant increase with a small effect size (SMD= 0.217) in AD patients compared to controls. Kendall’s tau and Egger’s regression (**Table 3**) indicate possible bias with 4 studies missing on the right side. After imputing these data, the difference was no longer significant (SMD= 0.147; 95%CI: -0.001; 0.294).

#### The 3HK/KYN ratio

ESF, Figure 4, and **Table 2** show that there is no significant difference in the 3HK/KYN between AD patients and healthy controls. **Table 3** revealed a possible bias with 3 studies missing on the right side.

**Figure 4:**
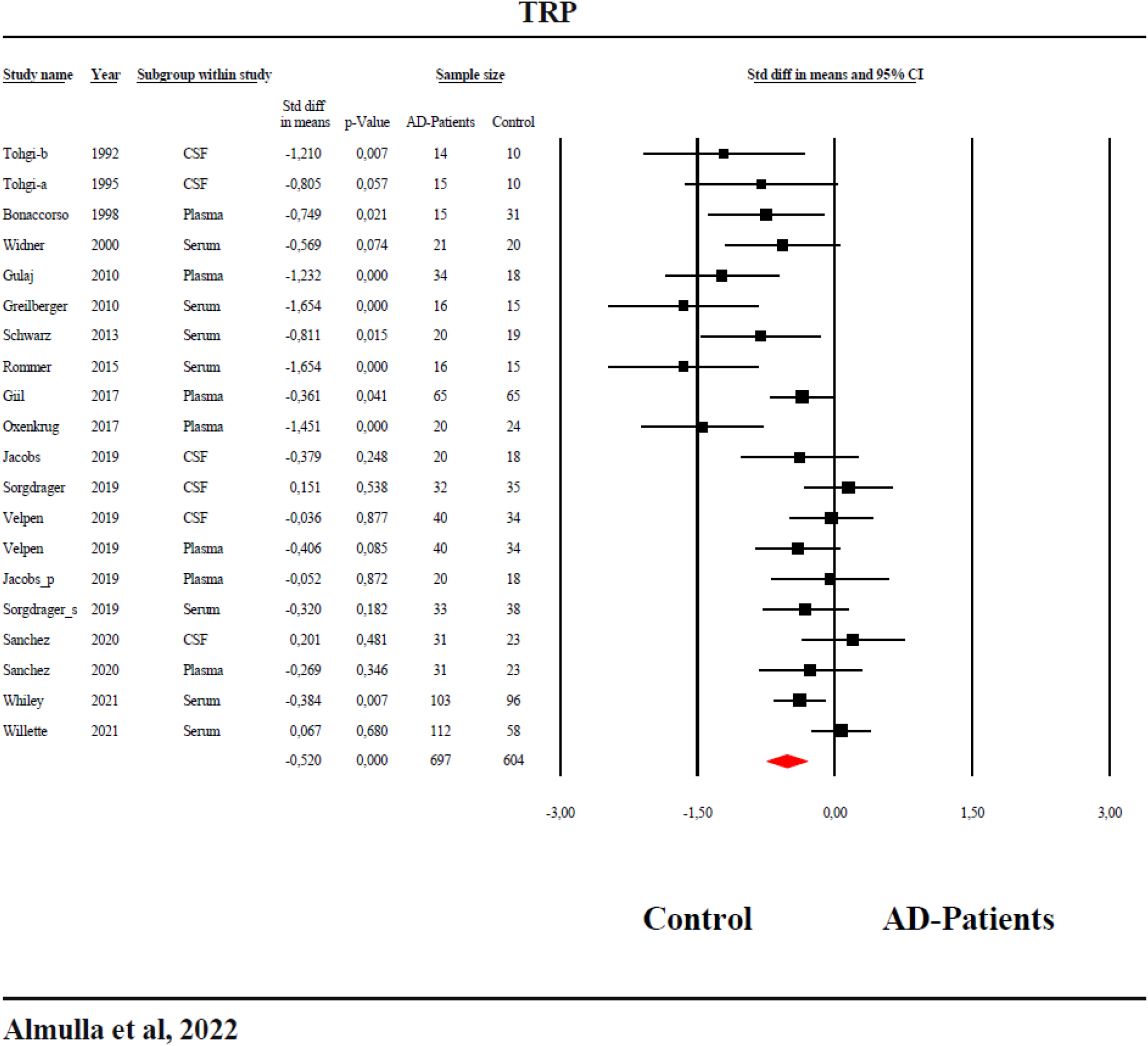
Forest plot with results of the meta-analysis performed on 20 studies reporting on tryptophan (TRP) in Alzheimer disease.

#### The(KYN+3HK+XA+QA+PA)/(KA+AA) ratio

There was no significant difference between AD patients and healthy controls in the neurotoxicity ratio (see **Table 2** and ESF, Figure 5). Publication bias was found with 4 studies missing on the left side.

### TRP and solitary TRYCATs

**Table 1** shows the distributions of the 95% CI. **Table 2** shows that TRP was significantly decreased with a moderate effect size in patients with AD compared to healthy controls. Kendall’s tau and Egger’s regression indicate a significant bias with 2 studies missing on the right. The adjusted estimate point was still significant (**Table 3**).

**Table 2** shows that there was no significant difference in KYN between AD patients and healthy controls (see **Table 2** and ESF, Figure 6). AD patients showed a significant decrease in KA levels with a small effect size, however, there was a significant difference (*p*=0.043) between the central and peripheral results. In plasma, a significant decrease in KA was detected, whereas in CNS and serum no significant alterations were found. ESF, Figure 7 shows the forest plot of KA results. The results of 3HK studies (**Table 1** and **Table 2**) show no significant differences in 3HK between AD and controls. There were no significant differences in QA and AA between patients and controls, although there was a trend towards lowered AA levels in AD. XA results were pooled from 5 studies.

**Table 2** and ESF, Figure 9 show a significant difference in XA between AD patients and healthy controls. After imputing 2 missing studies on the right, the SMD remained significant.

### Meta-regression analysis

Meta-regression results are presented in ESF, Table 6. Briefly, it shows that gender, whether male/female or both, significantly affects most of the outcome data, namely KYN/TRP, (KA+KYN)/TRP, KA/KYN, (KYN+3HK+XA+QA+PA)/(KA+AA), TRP, and KYN. Age was found to influence the results of KYN/TRP, 3HK/KYN, 3HK, and TRP. The sample size affected the outcome of KYN/TRP, (KA+KYN)/TRP, and TRP. The other confounders, such as latitude, MMSE scores. etc., exerted some effect on single or multiple outcomes.

## Discussion

The first major finding of this systematic review and meta-analysis is that both ratios reflecting the KYN/TRP ratio were significantly higher in AD than controls and, after adjusting for possible bias, all significance disappeared. Moreover, our meta-analysis did not show any changes in KYN in AD, whilst TRP levels were significantly decreased in AD patients. These results indicate normal IDO activity in AD patients and lowered availability of TRP, which may be explained by AD-associated reductions in levels of albumin that bind TRP (Maes, DeVos et al. 1999, Kim, Byun et al. 2020; Maes, Wauters et al. 1996). Furthermore, TRP availability to the brain is an age-sensitive process (Bonaccorso, Lin et al. 1998), which may contribute to low CNS levels of TRP in AD. In theory, reduced levels of TRP in AD may be due to malnutrition associated with cognitive dysfunction of AD patients (Giil, Midttun et al. 2017). Lowered levels of TRP in AD could be associated with increased behavioral and psychological symptoms of dementia (BPSD) including depression (Hood, Bell et al. 2005, Ruhé, Mason et al. 2007). Furthermore, TRP has neuroprotective effects and thus decreases may further aggravate neurodegenerative processes (Del Angel-Meza, Dávalos-Marín et al. 2011).

The second major finding is that we detected a significant increase in the KA/KYN ratio in the CNS of AD patients but not in peripheral blood. In contrast, the KA/(KYN+TRP) ratio was significantly increased with a very small effect size in the total study group. Thus, these results suggest a mild activation of KAT enzyme in the CNS of patients with AD. Previous papers showed no changes in peripheral KAT activity (Hartai, Juhasz et al. 2007), although centrally KAT was activated (Baran, Jellinger et al. 1999). However, our findings revealed a significant decrease in plasma KA levels in AD without changes in CNS and serum levels. In line with our results, Hartai et al reported lowered levels of KA in CSF and plasma (Hartai, Juhasz et al. 2007). In addition, no change in CSF KA levels was found in AD (Wennstrom, Nielsen et al. 2014). Therefore, it is likely that the lowered KA production is the consequence of lowered levels of KYN and KAT activity as well. Furthermore, KA formation is influenced by aberrations in energy metabolism (Hodgkins and Schwarcz 1998), an early hallmark of AD (Hartai, Juhasz et al. 2007).

KMO enzyme activity, as reflected by the 3HK/KYN ratio, was not significantly changed in AD. Apart from this, our results also indicate no significant change in overall 3HK levels, confirming our KMO results. Previous studies reported no changes in 3HK levels in AD patients compared to controls, in either central or peripheral levels (Baran, Jellinger et al. 1999, Gulaj, Pawlak et al. 2010, Jacobs, Lim et al. 2019, Sorgdrager, Vermeiren et al. 2019). KMO enzyme requires NADPH as a coenzyme (Hughes, Guner et al. 2022) and, therefore, the overactivity of NADPH oxidase enzyme consumes NADPH in AD patients (Fragoso-Morales, Correa-Basurto et al. 2021), which may cause attenuated KMO enzyme activity.

The fourth major finding of the present study is that there was no significant difference in the TRYCAT neurotoxicity (KYN+3HK+XA+QA+PA) / (KA+AA) ratio between AD patients and controls. Furthermore, our results showed a significant decrease in XA levels, one of the neurotoxic TRYCATs, in AD versus controls. Previous studies reported low XA levels in AD patients (Giil, Midttun et al. 2017, Sorgdrager, Vermeiren et al. 2019). The decreased XA in AD may in part be ascribed to lowered levels of 3HK and lowered TRYCAT pathway activity because of lowered TRP levels. In addition, XA may decrease in response to high levels of free radicals which are detected in AD (Giil, Midttun et al. 2017).

We also found that QA was not significantly altered in AD patients compared to controls. For example, Greilberger et al and Schwarz reported no changes in QA levels (Greilberger, Fuchs et al. 2010, Schwarz, Guillemin et al. 2013). It is hypothesized that in AD, QA is potentially implicated in increased tau protein phosphorylation and contributes to the neurofibrillary tangles (Rahman, Ting et al. 2009, Lugo-Huitrón, Ugalde Muñiz et al. 2013). However, QA is largely produced from 3HK by activated microglia in the CNS and no changes in 3HK could be detected in our meta-analysis. Sorgdrager et al suggested that serum and CSF levels of QA in neurodegenerative disease may accumulate in response to aging (Sorgdrager, Vermeiren et al. 2019). Regardless, our results indicate that neurotoxicity due to increased levels of the neurotoxic TRYCATs does not play a role in AD.

Finally, AA levels were significantly decreased in AD patients compared with controls, as previously reported in an early study (Oxenkrug, van der Hart et al. 2017). AA formation is regulated by activity of kynureninase, which catalyzes AA formation from KYN, an immediate substrate for biosynthesis of AA, 3HK and KYNA. Notably, kynureninase affinity to KYN is much lower than to KMO and KAT, and under physiological conditions KYN is mainly converted into 3HK and KYNA rather than into AA (Moroni 1999). Up-regulation of KMO, which catalyzes KYN formation into 3HK, might decrease the availability of KYN as a substrate for kynureninase and, consequently, down-regulate AA production. Elevation of 3HK serum levels might provide a mechanistical explanation of down-regulated AA formation (Oxenkrug, van der Hart et al. 2017). However, the present meta-analysis reveals no significant differences in KYN and 3HK between AD and controls that might explain decreased plasma/serum AA levels. On the other hand, elevated plasma AA levels was the most powerful predictor of future development of incident dementia (Chouraki, Preis et al. 2017). Higher AA (and KYN) plasma levels were observed in high neocortical amyloid-beta load (NAL) than in noo-NAL female (but not male) patients prior to development of cognitive impairment, and AA (and KYN) levels individually and jointly predicted development of NAL(Chatterjee, Goozee et al. 2018). Notably, the alternative to IDO enzyme that catalyzes TRP conversion into KYN is tryptophan 2,3-dioxygenase 2 (TDO), which is activated by cortisol. Correlations between post-dexamethasone cortisol levels were observed in female, but not male, AD patients (Oxenkrug, Gurevich et al. 1989).

### Limitations

Limitations regarding the current systematic review and meta-analysis are as follows: First, most included studies did not delineate the exact stage of AD, and we were, therefore, not able to examine the stage-related changes in TRYCATs levels. Second, there was a lack of information on the patients’ treatments which might affect TRYCATs levels. Third, the lack of postmortem studies and small sample sizes concerning CSF studies restrict our ability to draw conclusions on the TRYCAT status in the brain. Lastly, most studies did not report cognitive impairments and psychiatric comorbidities, which may further change TRYCAT pathway activity (Almulla, Vasupanrajit et al. 2021).

## Conclusions

Figure 1. summarizes the results of the present study. Our findings indicate that the availability of TRP is reduced in patients with AD, although there are no signs that IDO is activated. Lowered TRP availability to the brain may be explained by lowered plasma albumin in AD. Moreover, there is a trend towards lowered AA, KA, and XA in AD, which may in part be explained by lowered levels of TRP. Since the composite TRYCAT neurotoxicity index was not increased in AD, we conclude that TRYCATs may not contribute to the neurotoxic pathophysiology of AD.

## Data Availability

On a reasonable request and after the authors have fully used the dataset, the employed dataset (excel file) developed for this meta-analysis will be accessible from MM.

## Declaration of Competing Interests

The authors declare that this work is free from any conflict of interest.

## Ethical approval and consent to participate

Not applicable.

## Consent for publication

Not applicable.

## Funding

The study was funded by the C2F program, Chulalongkorn University, Thailand.

## Author’s contributions

All the authors contributed to the writing of this work. AA and MM designed the study. AA and TS collected the data. AA and MM performed the statistical analysis.

## Acknowledgments

Not applicable.

## Notes

### Competing Interest Statement

The authors have declared no competing interest.

