## supplementary file for "The tryptophan catabolite or kynurenine pathway in Alzheimer’s disease: a systematic review and meta-analysis"

SHORT TITLE: Kynurenine pathway in Alzheimer's disease

Abbas F. Almulla, Ph.D.[a,b] , Thitiporn Supasitthumrong, M.D. .[a], Arisara Amrapala, MS.c[a], Chavit Tunvirachaisakul, M.D., Ph.D.[a], Al-Karrar Kais Abdul Jaleel, MS.c[b], Gregory Oxenkrug, M.D., Ph.D.[c], Hussein K. Al-Hakeim, Ph.D.[d], Michael Maes, M.D., Ph.D.[a,e,f]

[a] Department of Psychiatry, Faculty of Medicine, Chulalongkorn University, Bangkok, Thailand.

[b] Medical Laboratory Technology Department, College of Medical Technology, The Islamic University, Najaf, Iraq.

[c] Department of Psychiatry, Tufts University School of Medicine and Tufts Medical Center, Boston, MA 02111, USA.

[d] Department of Chemistry, College of Science, University of Kufa, Kufa, Iraq.

[e] Department of Psychiatry, Medical University of Plovdiv, Plovdiv, Bulgaria.

[f] Department of Psychiatry, IMPACT Strategic Research Centre, Deakin University, Geelong, Victoria, Australia.

**Corresponding author:**

Prof. Dr Michael Maes, M.D., Ph.D.

Department of Psychiatry

Faculty of Medicine, Chulalongkorn University

Bangkok, 10330

Thailand

E-mail addresses:

#### ESF, Table 1: Search sentences and terms were used in each database

| Database Name | Search Sentence | No. of Articles |
|---|---|---|
| **PubMed/Medline** | ((((((((((((Alzheimer's disease and TRYCATs)* OR (Dementia and kynurenine)) OR (Alzheimer's disease and kynurenic acid)) OR (Alzheimer's disease and quinolinic acid)) OR (Alzheimer's disease and Picolinic acid)) OR (Alzheimer's disease and 3-hydroxyanthranilic acid)) OR (Alzheimer's disease and xanthurenic acid)) OR (Alzheimer's disease and 3-hydroxykynurenine)) OR (Alzheimer's disease and Anthranilic acid)) OR (Alzheimer's disease and formyl kynurenine)) OR (Alzheimer's disease and l-tryptophan)) OR (Alzheimer's disease and tryptophan catabolites)) OR (TRYCATs and Alzheimer's disease)) OR (TRYCATs and AD) | **707** |
| | ((((((((((TRYCAT pathway and Dementia) OR (TRYCATs and mild cognitive impairment)) OR (TRYCAT pathway and Alzheimer's disease)) OR (Kynurenine pathway and Alzheimer's disease)) OR (IDO and Alzheimer's disease)) OR (kynurenine and dementia)) OR (kynurenic acid and dementia)) OR (3-Hydroxykynurenine and dementia)) OR (xanthurenic acid and dementia)) OR (3-Hydroxyanthranilic acid and dementia)) OR (Quinolinic acid and dementia)) OR (Picolinic acid and dementia) | **387** |
| | (((((((((Alzheimer's disease and IDO enzyme)* OR (Alzheimer's disease and TDO enzyme)) OR (Alzheimer's disease and KAT enzyme)) OR (Alzheimer's disease and KMO enzyme)) OR (Alzheimer's disease and Kynureninase enzyme)) OR (SARS-Dementia and IDO enzyme)) OR (Dementia and KAT enzyme)) OR (Dementia and Kynurenine pathway)) OR (Dementia and Kynureninase enzyme)) OR (Dementia and TDO) | **2075** |
| **Google Scholar** | (((((Alzheimer's disease* and IDO activation) OR (TDO activation)) OR (Kynurenine pathway)) OR (KMO activation)) OR (Tryptophan degradation)) AND (Decreased Tryptophan) | **3390** |
| | (((((Alzheimer's disease* and kynurenine pathway) OR (TRYCAT pathway)) OR (Tryptophan)) OR (kynurenine)) OR (kynurenic acid)) AND (3- | **2220** |

| | | |
|---|---|---|
| | Hydroxykynurenine) OR (Xanthurenic acid)) OR (Quinolinic acid)) OR (Picolinic acid)) OR (Anthranilic acid)) OR (3-Hydroxyanthranilic acid)) | |
| **Web of Science** | (((((((((ALL=(Alzheimer's disease and Tryptophan)) OR ALL=(Alzheimer's disease and Kynurenine pathway)) OR ALL=(Alzheimer's disease and kynurenine)) OR ALL=(Alzheimer's disease and kynurenic acid)) OR ALL=(Alzheimer's disease and 3-Hydroxykynurenine)) OR ALL=(Alzheimer's disease and Xanthurenic acid )) OR ALL=(Alzheimer's disease and Anthranilic acid)) OR ALL=(Alzheimer's disease and Quinolinic acid)) OR ALL=(Alzheimer's disease and 3-Hydroxyanthranilic acid)) OR ALL=(Alzheimer's disease and Picolinic acid) | **630** |
| | (((((((((ALL=(Alzheimer's disease and IDO enzyme)) OR ALL=(Alzheimer's disease and KAT enzyme )) OR ALL=(Alzheimer's disease and KMO enzyme)) OR ALL=(Alzheimer's disease and TDO enzyme)) OR ALL=(Alzheimer's disease and Kynureninase enzyme)) OR ALL=(Dementia and kynurenine pathway)) OR ALL=(Dementia and TRYCATs)) OR ALL=(Dementia and TRYCAT pathway)) OR ALL=(Dementia and tryptophan )) OR ALL=(Dementia and kynurenine) | **395** |
| **SciFinder** | Alzheimer's disease and kynurenine pathway, Alzheimer's disease and tryptophan catabolism pathway, Alzheimer's disease and TRYCATs, Alzheimer's disease and kynurenine, Alzheimer's disease and tryptophan, Alzheimer's disease kynurenic acid, Dementia and kynurenine pathway | **70** |

**EFS, Table 2:** Studies excluded from the meta-analysis but included in the systematic review.

| Authors, year | Reason why excluded from the meta-analysis |
|---|---|
| Porter, Lunn et al. 2003 | The author did not show a quantitive data |
| Guillemin, Brew et al. 2005 | No data for measured analytes were presented |
| Trushina, Dutta et al. 2013 | No mean (SD) for measured analytes |
| Bonda, Mailankot et al. 2010 | The article included antibody against TRYCATs |
| Porter, Lunn et al. 2003 | No data for the baseline levels |

**ESF, Table 3. Immune cofounder's scale (ICS) applied from Andrés-Rodríguez, et al., 2019**

| | |
|---|---|
| **Methodological quality of the study** | |
| **1** | Study sample ≥ 128 participants including patients and controls (1= Yes, 0 = No) |
| **2** | Did the study control the results for potential confounders (e.g., age, BMI, gender, race)? (1= Yes, 0 = No) |
| **3** | Were participants with schizophrenia and controls age- and-gender-matched or was there a statistical control? (1= Yes, 0 = No) |
| **4** | Was the time of sample collection specified (e.g., morning vs. evening)? (1= Yes, 0 = No) |
| **5** | Were participants with Alzheimer disease free of immunomodulatory drugs including anti-cytokines, glucocorticoids, immunoglobulins, and immunosuppressants, or was there a medication washout period or was drug intake statistically controlled for? (1= Yes, 0 = No) |
| **6** | Were participants with schizophrenia free of nervous system drugs or were the data statistically controlled for? (1= Yes, 0 = No) |
| **7** | Reporting either the manufacturer of the test or detection limit and coefficients of variation (1= Yes, 0 = No) |
| **8** | Reporting how data under detection limit were handled (1 = Yes, 0 = No) |
| **9** | Reporting % of the sample under detection limit (1=Yes, 0= No) |
| **10** | Reporting blood fraction (serum, plasma, culture supernatant or whole blood) (1= Yes, 0 = No) |
| | **Total quality score (10 points)** |
| **Biomarker confounders red points**<br>*The red points should not be given if the item is statistically controlled for* | |
| **1** | 3 red points for comorbid illnesses such as autoimmune disorders & other immune disorders including rheumatoid arthritis, psoriasis, inflammatory bowel disease, chronic obstructive pulmonary disease, multiple sclerosis |
| **2** | 3 red points for use of recreational drugs such as methamphetamine or opioids |
| **3** | 2 red points when groups were not matched for age |
| **4** | 2 red points when groups were not matched for sex |

| | |
|---|---|
| **5** | 2 red points for medication use as for example immunomodulators |
| **6** | 2 red points for early traumatic life events |
| **7** | 2 red points for shift work and primary sleep disorders |
| **8** | 1.5 red points for use of neuroleptics |
| **9** | 1 red point for more common systemic immune disorders including diabetes type 1/2, essential hypertension, metabolic syndrome |
| **10** | 1 red point for not fasting (8 hours before blood extraction) |
| **11** | 1 red point for use of omega-3 and antioxidant supplements |
| **12** | 1 red point when data were not controlled for body mass index |
| **13** | 1 red point when data were not controlled for physical activity or sedentary life |
| **14** | 1 red point when data were not controlled for smoking |
| **15** | 1 red point for use of oral contraceptives or NSAIDs |
| **16** | 0.5 red points when data were not controlled for ethnicity in countries such as US, Brazil |
| **17** | 0.5 red points when data were not controlled for seasonality |
| **18** | 0.5 red points when data were not controlled for diurnal variation (8-10 a.m. versus all other time points) |
| | **Total red point score (26 points)** |

**ESF, Table 4. PRISMA checklist**

| Section/topic | # | Checklist item | Reported on page # |
|---|---|---|---|
| **TITLE** | | | |
| Title | 1 | Identify the report as a systematic review, meta-analysis, or both. | 1 |
| **ABSTRACT** | | | |
| Structured summary | 2 | Provide a structured summary including, as applicable: background; objectives; data sources; study eligibility criteria, participants, and interventions; study appraisal and synthesis methods; results; limitations; conclusions and implications of key findings; systematic review registration number. | 3 |
| **INTRODUCTION** | | | |
| Rationale | 3 | Describe the rationale for the review in the context of what is already known. | 5 |
| Objectives | 4 | Provide an explicit statement of questions being addressed with reference to participants, interventions, comparisons, outcomes, and study design (PICOS). | 9 |
| **METHODS** | | | |
| Protocol and registration | 5 | Indicate if a review protocol exists, if and where it can be accessed (e.g., Web address), and, if available, provide registration information including registration number. | 10 |
| Eligibility criteria | 6 | Specify study characteristics (e.g., PICOS, length of follow-up) and report characteristics (e.g., years considered, language, publication status) used as criteria for eligibility, giving rationale. | 11 |
| Information sources | 7 | Describe all information sources (e.g., databases with dates of coverage, contact with study authors to identify additional studies) in the search and date last searched. | 11 |
| Search | 8 | Present full electronic search strategy for at least one database, including any limits used, such that it could be repeated. | 11 |
| Study selection | 9 | State the process for selecting studies (i.e., screening, eligibility, included in systematic review, and, if applicable, included in the meta-analysis). | 11 |
| Data collection process | 10 | Describe method of data extraction from reports (e.g., piloted forms, independently, in duplicate) and any processes for obtaining and confirming data from investigators. | 11 |
| Data items | 11 | List and define all variables for which data were sought (e.g., PICOS, funding sources) and any assumptions and simplifications made. | 11 |
| Risk of bias in individual studies | 12 | Describe methods used for assessing risk of bias of individual studies (including specification of whether this | 12 |

| | | | |
|---|---|---|---|
| | | was done at the study or outcome level), and how this information is to be used in any data synthesis. | |
| Summary measures | 13 | State the principal summary measures (e.g., risk ratio, difference in means). | 12 |
| Synthesis of results | 14 | Describe the methods of handling data and combining results of studies, if done, including measures of consistency (e.g., $I^2$) for each meta-analysis. | 12 |
| Risk of bias across studies | 15 | Specify any assessment of risk of bias that may affect the cumulative evidence (e.g., publication bias, selective reporting within studies). | 13 |
| Additional analyses | 16 | Describe methods of additional analyses (e.g., sensitivity or subgroup analyses, meta-regression), if done, indicating which were pre-specified. | 14 |
| **RESULTS** | | | |
| Study selection | 17 | Give numbers of studies screened, assessed for eligibility, and included in the review, with reasons for exclusions at each stage, ideally with a flow diagram. | 15 |
| Study characteristics | 18 | For each study, present characteristics for which data were extracted (e.g., study size, PICOS, follow-up period) and provide the citations. | 15 |
| Risk of bias within studies | 19 | Present data on risk of bias of each study and, if available, any outcome level assessment (see item 12). | ESF, 8, table 6 |
| Results of individual studies | 20 | For all outcomes considered (benefits or harms), present, for each study: (a) simple summary data for each intervention group (b) effect estimates and confidence intervals, ideally with a forest plot. | ESF, 7, table 5 |
| Synthesis of results | 21 | Present results of each meta-analysis done, including confidence intervals and measures of consistency. | 45, table 2 |
| Risk of bias across studies | 22 | Present results of any assessment of risk of bias across studies (see Item 15). | ESF, 8, table 6 |
| Additional analysis | 23 | Give results of additional analyses, if done (e.g., sensitivity or subgroup analyses, meta-regression [see Item 16]). | ESF, 9, table 7 |
| **DISCUSSION** | | | |
| Summary of evidence | 24 | Summarize the main findings including the strength of evidence for each main outcome; consider their relevance to key groups (e.g., healthcare providers, users, and policy makers). | 21-25 |
| Limitations | 25 | Discuss limitations at study and outcome level (e.g., risk of bias), and at review-level (e.g., incomplete retrieval of identified research, reporting bias). | 26 |
| Conclusions | 26 | Provide a general interpretation of the results in the context of other evidence, and implications for future research. | 26, Figure 6 |
| **FUNDING** | | | |
| Funding | 27 | Describe sources of funding for the systematic review and other support (e.g., supply of data); role of funders for the | 27 |

| | | |
|---|---|---|
| | | systematic review. |

ESF, Table 5: Characteristics of Studies which included in the systematic reviews and meta-analysis

| NO | Authors, years | Setting | Type of case | Type of Control | Sample Size | | | Age | | Assessed Biomarkers | Specimen | Method | Quality score | Red point score | Findnigs in AD patients compared to HC |
|---|---|---|---|---|---|---|---|---|---|---|---|---|---|---|---|
| | | | | | Cases (M/F) | Control M/F | Total M/F | Case-Mean (SD) | Control-Mean(SD) | | | | | | |
| 1 | Oxenkrug, van der Hart et al. 2017 | USA | AD | HC | 20 (12/8) | 24 (12/12) | 44 (24/20) | - | - | 3HK,AA,KA,KYN,TRP,XA | Plasma | HPLC-MS | 4 | 13.5 | Low TRP, AA,KA High KYN,3HK, XA |
| 2 | Giil, Midttun et al. 2017 | Norway | AD | HC | 65 (37/28) | 65 (32/33) | 130 (69/51) | 74,3(15,1) | 81,6(8,6) | 3HK, AA, KA,KYN, QA, TRP, XA | Plasma | LC-MS/MS | 6 | 12,5 | Low TRP, KYN, 3HK, KA, XA, AA, QA |
| 3 | Rommer, Fuchs et al. 2016 | Austria | AD | HC | 16 (7/9) | 15 (4/11) | 31 (11/20) | 63,3(13,7) | 62,8(3,6) | TRP, KYN | Serum | LC | 7,5 | 12 | Low TRP High KYN |
| 4 | Wennstrom, Nielsen et al. 2014 | Sweden | AD | HC | 19 (9/10) | 20 (10/10) | 39 (10/20) | 75,3(5,6) | 77(10,3) | KA | CSF | HPLC | 4,5 | 11 | Low KA |
| 5 | Schwarz, Guillemin et al. 2013 | Germany | AD | HC | 20 (4/16) | 19 (11/8) | 39 (15/24) | 74(7,6) | 59,5(10,2) | 3HK, KA, KYN, QA, TRP | Serum | HPLC | 5,5 | 12,5 | Low TRP, KA, QA High KYN, 3HK, PA |
| 6 | Gulaj, Pawlak et al. 2010 | Poland | AD | HC | 34 (10/24) | 18 (5/13) | 52 (15/37) | 78,8(5,6) | 76,1(7,3) | 3HK, AA, KA, KYN, QA, TRP | Plasma | HPLC | 8 | 10,5 | Low ( TRP, 3HK,AA,KA) High (QA,KYN) |
| 7 | Greilberger, Fuchs et al. 2010 | Austria | AD | HC | 16 (7/9) | 15 (4/11) | 31 (11/20) | 63,3(13,7) | 62,8(3,6) | TRP, KYN | Serum | HPLC | 4,5 | 10 | Low TRP High KYN |
| 8 | Hartai, Juhasz et al. 2007 | Hungary | AD | HC | 28 (6/22) | 31 (10/21) | 59 (16/43) | 77(6.3) | 73(8.3) | KYN, KA | Plasma | HPLC | 4,5 | 11 | Low KA, KYN |
| 9 | Widner, Leblhuber et al. 2000 | Austria | AD | HC | 21 (6/15) | 20 (10/10) | 41 (16/25) | 74,4(5,4) | 73,4(7,4) | TRP, KYN | Serum | HPLC | 5 | 10 | Low TRP High KYN |
| 10 | Baran, Jellinger et al. 1999 | Austria | AD | HC | 11 (2/9) | 13 (7/6) | 24 (9/15) | 81(6,3) | 80,1(7,9) | 3HK, KA, KYN, | Brain tissue | HPLC | 5 | 11 | Low KYN, 3HK High KA |
| 11 | Bonaccorso, Lin et al. 1998 | Belgium | AD | HC | 15 (3/12) | 31 (14/17) | 46 (17/29) | 78,4(10,3) | 55,2(22,2) | TRP | Plasma | HPLC | 5,5 | 10 | Low TRP |
| 12 | Tohgi, Abe et al. 1995 | Japan | AD | HC | 15 | 10 | 25 | 68(6) | 69(6) | TRP, KYN, 3HK | CSF | HPLC | 5,5 | 10 | Low TRP, KYN, 3HK |
| 13 | Tohgi, Abe et al. 1992 | Japan | AD | HC | 14 | 10 | 24 | 68,4(10,1) | 68,5(6,1) | TRP, KYN, 3HK | CSF | HPLC | 5,5 | 10 | Low TRP, KYN, 3HK |
| 14 | Willette, Pappas et al. 2021 | USA | AD | HC | 112 (65/47) | 58 (30/28) | 170 (95/75) | 74,8(8) | 75,1(5,7) | TRP, KYN | Serum | LC-MS | 9 | 10,5 | High TRP Low KYN |
| 15 | Whiley, Chappell et al. 2021 | UK | AD | CN | 103 (50/53) | 86 (44/42) | 189 (94/95) | 76,9(5,8) | 75,9(5,6) | TRP, KYN, XA | Serum | UHPLC -MS/MS | 5 | 11 | Low TRP, KYN, XA |

| | | | | | | | | | | | | | | | |
|---|---|---|---|---|---|---|---|---|---|---|---|---|---|---|---|
| 16 | Gonzalez-Sanchez, Jimenez et al. 2020 | Spain | AD | HC | 20,41 (7/13), (19/22) | 23 (15/8) | 43,64 (22/21), (34/30) | 71,9(8,1) | 64,7(10,8) | TRP, KA | Plasma and CSF | ELISA | 7,5 | 12,5 | High c(TRP, KA) Low p(TRP) High p(KA) |
| 17 | van der Velpen, Teav et al. 2019 | Switzerland | AD | HC | 40 (16/24) | 34 (11/23) | 74 (27/47) | 74.8(6.3) | 65.3(6.1) | KA, QA, TRP | Plasma and CSF | LC-MS | 6 | 11 | Low TRP, High KA, QA |
| 18 | Sorgdrager, Vermeiren et al. 2019 | Belgium | AD | HC | 33 (15/18) | 39 (18/21) | 72 (33/39) | 73,7(6) | 71,3(10,7) | 3HK, KYN, KA, QA, TRP, XA | Serum and CSF | UPLC | 4 | 12.5 | Low s(TRP, 3HK, XA, QA, KA) High sKYN Low c(3HK,KA,XA,QA) High c(TRP, KYN) |
| 19 | Jacobs, Lim et al. 2019 | Sweden | AD | HC | 20 (9/11) | 18 (15/3) | 38 (24/14) | 77,9(7,5) | 73,1(7,9 | TRP, KYN, KA, 3HK, 3HA, AA, PA, QA | Serum and CSF | UHPLC | 2,5 | 12,5 | Low s(TRP,KA) High c(KYN,3HK,AA,PA,QA) |

**Table 4: Results of Meta-regression**

| Variables | No. of Studies | Covariates | 1-sided p-value | Z-Value |
|---|---|---|---|---|
| KYN/TRP | 20 | Age | 0.043 | -1.71 |
| | | No. of cases | 0.027 | -1.92 |
| | | Sample size | 0.018 | -2.08 |
| | 18 | Male gender | 0.026 | -1.93 |
| | | Female gender | 0.013 | -2.20 |
| | 13 | MMSE | 0.031 | 1.87 |
| | 21 | Central-Peripheral | 0.024 | 1.98 |
| (KA+KYN)/TRP | 19 | Male gender | 0.043 | -1.71 |
| | | Female gender | 0.023 | -1.99 |
| | 21 | No. of cases | 0.038 | -1.77 |
| | | Sample size | 0.026 | -1.93 |
| KA/KYN | 17 | Male gender | 0.035 | 1.80 |
| | | Female gender | 0.010 | 2.30 |
| | 21 | Central-Peripheral | 0.003 | -2.67 |
| 3HK/KYN | 16 | Age | 0.012 | 2.23 |
| (KYN+3HK+XA+QA+PA)/(KA+AA) | 20 | Central-Peripheral | 0.001 | 3.10 |
| | 17 | Male gender | 0.043 | -1.71 |
| | | Female gender | 0.037 | -1.78 |
| 3HK | 11 | Age | 0.014 | 2.18 |
| | | Central-Peripheral | 0.009 | 2.34 |
| KA | 14 | Central-Peripheral | 0.007 | -2.45 |
| TRP | 17 | Age | 0.003 | 2.71 |
| | 18 | No. of cases | 0.009 | 2.35 |
| | | Sample size | 0.007 | 2.41 |
| | 16 | Male gender | 0.018 | 2.08 |
| | | Female gender | 0.022 | 2 |

| | | | | |
|---|---|---|---|---|
| KYN | 15 | Male gender | 0.023 | -1.98 |
| | | Female gender | 0.004 | -2.60 |

#### (KYN+KA)/TRP

| Study name | Year | Std diff in means | p-Value | Sample size: AD-Patients | Sample size: Control |
|---|---|---|---|---|---|
| Tohgi-b | 1992 | -0,350 | 0,461 | 14 | 10 |
| Tohgi-a | 1995 | 0,277 | 0,507 | 15 | 10 |
| Bonaccorso | 1998 | 0,749 | 0,021 | 15 | 31 |
| Baran | 1999 | 0,100 | 0,809 | 11 | 13 |
| Widner | 2000 | 0,434 | 0,170 | 21 | 20 |
| Hartai | 2007 | -0,575 | 0,031 | 28 | 31 |
| Gulaj | 2010 | 0,279 | 0,363 | 34 | 18 |
| Greilberger | 2010 | 1,038 | 0,008 | 16 | 15 |
| Schwarz | 2013 | 0,368 | 0,258 | 20 | 19 |
| Wennstrom | 2014 | -0,024 | 0,939 | 19 | 20 |
| Rommer | 2015 | 1,038 | 0,008 | 16 | 15 |
| Giil | 2017 | -0,101 | 0,568 | 65 | 65 |
| Oxenkrug | 2017 | 0,367 | 0,247 | 20 | 24 |
| Jacobs | 2019 | 0,464 | 0,163 | 20 | 18 |
| Jacobs | 2019 | 0,089 | 0,785 | 20 | 18 |
| Sorgdrager | 2019 | -0,288 | 0,244 | 32 | 35 |
| Sorgdrager | 2019 | 0,044 | 0,855 | 33 | 38 |
| Velpen | 2019 | 0,160 | 0,495 | 40 | 34 |
| Velpen | 2019 | 0,406 | 0,085 | 40 | 34 |
| Sanchez | 2020 | 0,589 | 0,093 | 22 | 22 |
| Sanchez | 2020 | 0,171 | 0,603 | 22 | 22 |
| Whiley | 2021 | -0,039 | 0,787 | 103 | 96 |
| Willette | 2021 | -0,043 | 0,791 | 112 | 58 |
| | | 0,167 | 0,027 | 738 | 665 |

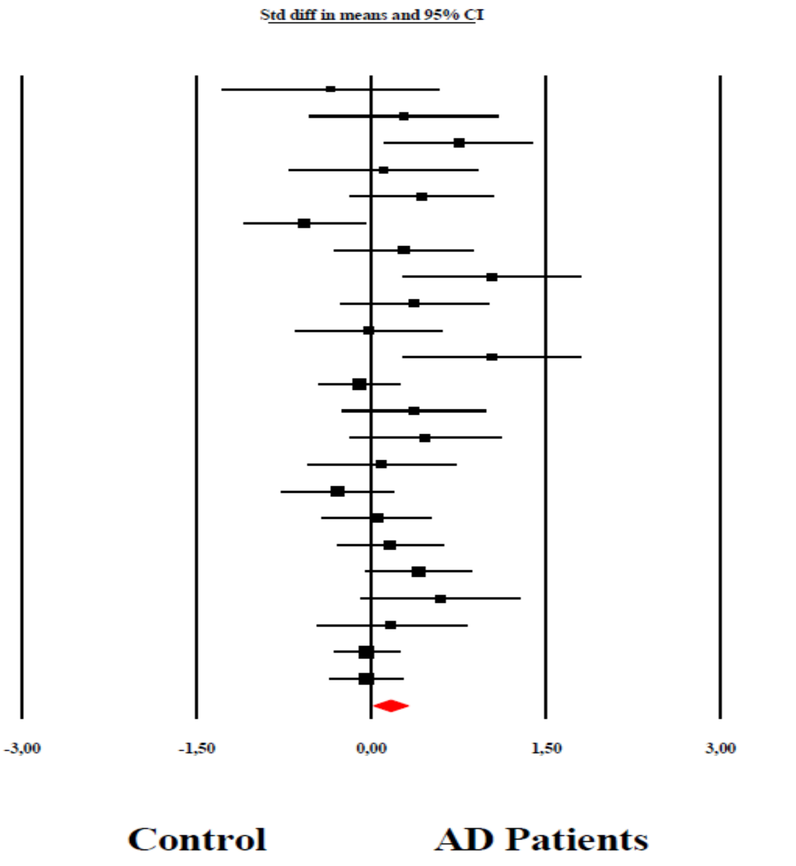

Almulla et al, 2022

**ESF, Figure 1:** The forest plot of kynurenine (KYN)+Kynurenic acid (KA)/ Tryptophan (TRP) ratio in patients with Alzheimer disease (AD) compared to healthy control reflecting IDO enzyme activity**.**

### KA/KYN

| Study name | Year | Subgroup within study | Std diff in means | p-Value | AD-Patients | Control |
|---|---|---|---|---|---|---|
| Baran | 1999 | CNS | 0,389 | 0,348 | 11 | 13 |
| Jacobs | 2019 | CNS | 0,465 | 0,164 | 20 | 18 |
| Sanchez | 2020 | CNS | 1,378 | 0,001 | 13 | 20 |
| Sorgdrager | 2019 | CNS | -0,336 | 0,176 | 32 | 35 |
| Tohgi-a | 1995 | CNS | 0,252 | 0,539 | 15 | 10 |
| Tohgi-b | 1992 | CNS | 1,909 | 0,000 | 14 | 10 |
| Velpen | 2019 | CNS | 0,283 | 0,227 | 40 | 34 |
| Wennstrom | 2014 | CNS | -0,024 | 0,939 | 19 | 20 |
| | | CNS | 0,466 | 0,042 | 164 | 160 |
| Giil | 2017 | Plasma | -0,032 | 0,856 | 65 | 65 |
| Gulaj | 2010 | Plasma | -0,793 | 0,009 | 34 | 18 |
| Hartai | 2007 | Plasma | -0,178 | 0,503 | 28 | 31 |
| Jacobs_p | 2019 | Plasma | -0,175 | 0,590 | 20 | 18 |
| Oxenkrug | 2017 | Plasma | -0,275 | 0,366 | 20 | 24 |
| Sanchez | 2020 | Plasma | 0,073 | 0,843 | 13 | 20 |
| | | Plasma | -0,198 | 0,082 | 180 | 176 |
| Greilberger | 2010 | Serum | -0,422 | 0,246 | 16 | 15 |
| Rommer | 2015 | Serum | -0,422 | 0,246 | 16 | 15 |
| Schwarz | 2013 | Serum | -0,161 | 0,616 | 20 | 19 |
| Sorgdrager_s | 2019 | Serum | -0,133 | 0,576 | 33 | 38 |
| Whiley | 2021 | Serum | 0,461 | 0,001 | 103 | 96 |
| Widner | 2000 | Serum | -0,300 | 0,340 | 21 | 20 |
| Willette | 2021 | Serum | 0,019 | 0,907 | 112 | 58 |
| | | Serum | -0,054 | 0,704 | 321 | 261 |
| | | Overall | -0,062 | 0,456 | 665 | 597 |

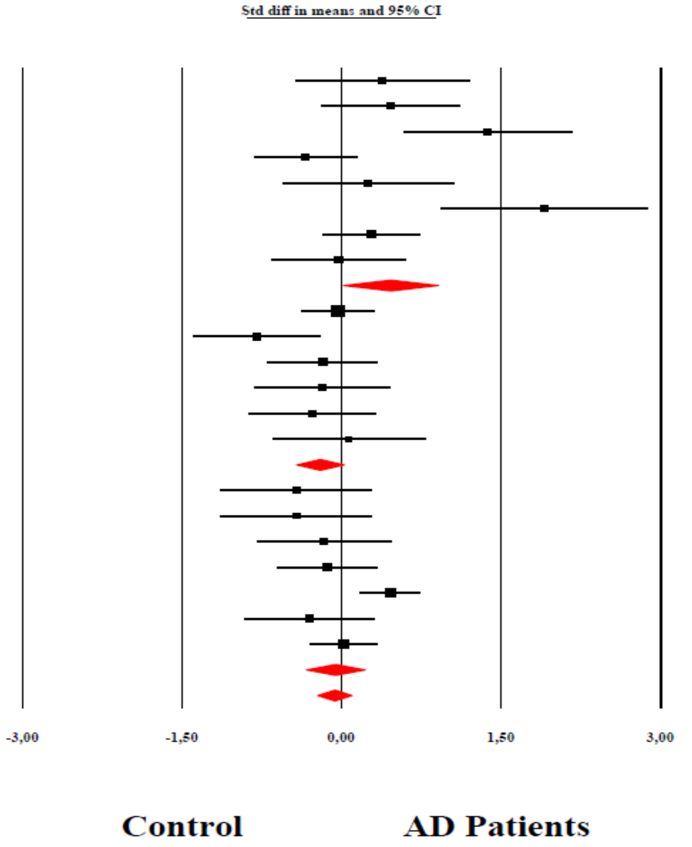

Almulla et al, 2022

**ESF, Figure 2:** The forest plot of Kynurenic acid (KA)/kynurenine (KYN)ratio in patients with Alzheimer disease (AD) compared to healthy control reflecting KAT enzyme activity**.**

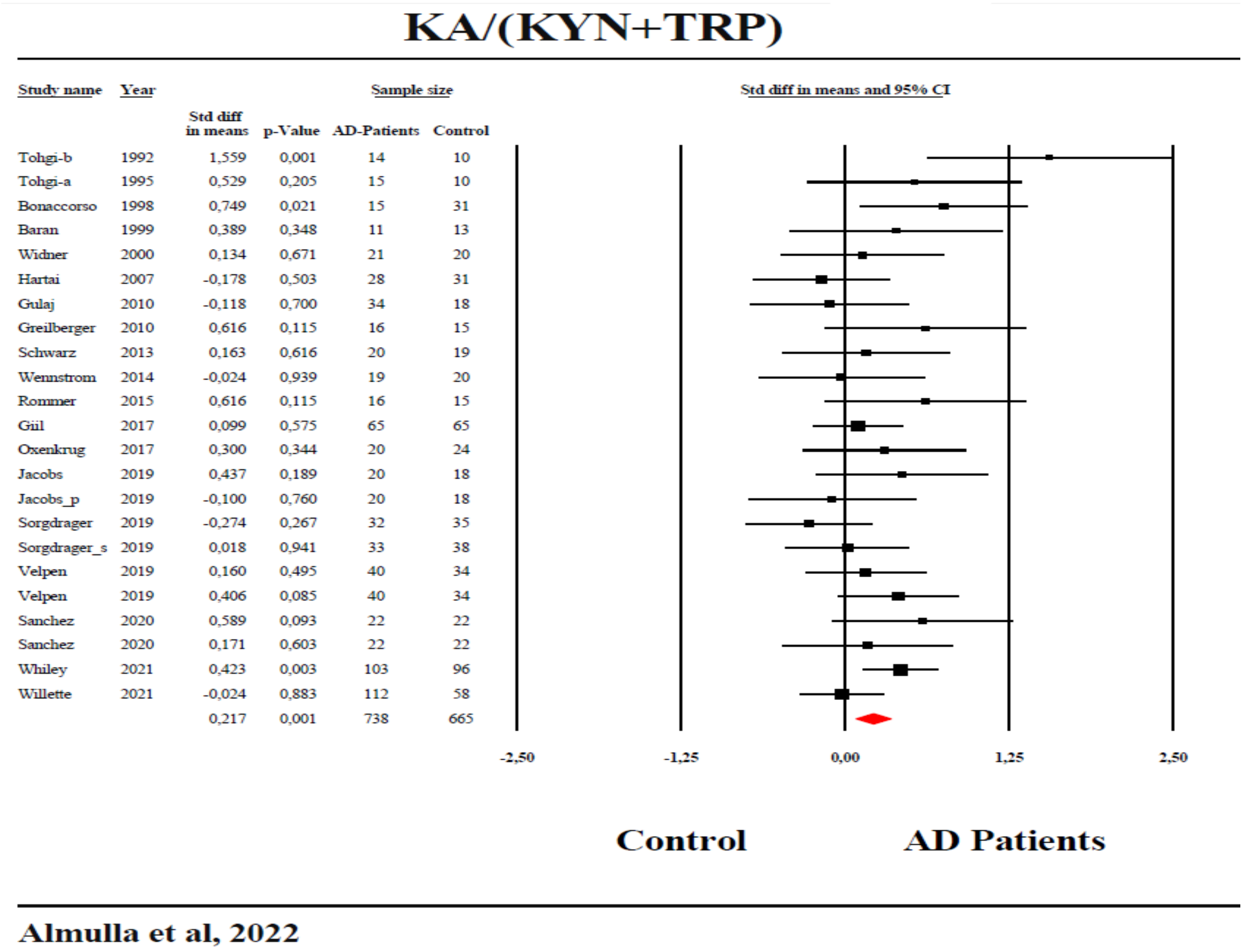

**ESF, Figure 3:** The forest plot of Kynurenic acid (KA)/kynurenine (KYN)+ Tryptophan (TRP) ratio in patients with Alzheimer disease (AD) compared to healthy control reflecting KAT enzyme activity.

#### 3HK/KYN

| Study name | Year | Std diff in means | p-Value | AD-Patients | Control |
|---|---|---|---|---|---|
| | | | | Sample size | |
| Tohgi-b | 1992 | -0,175 | 0,733 | 14 | 10 |
| Tohgi-a | 1995 | -0,559 | 0,195 | 15 | 10 |
| Baran | 1999 | -0,106 | 0,798 | 11 | 13 |
| Widner | 2000 | -0,300 | 0,340 | 21 | 20 |
| Hartai | 2007 | 0,396 | 0,132 | 28 | 31 |
| Gulaj | 2010 | -0,261 | 0,375 | 34 | 18 |
| Greilberger | 2010 | -0,422 | 0,246 | 16 | 15 |
| Schwarz | 2013 | 0,469 | 0,163 | 20 | 19 |
| Rommer | 2015 | -0,422 | 0,246 | 16 | 15 |
| Giil | 2017 | -0,090 | 0,612 | 65 | 65 |
| Oxenkrug | 2017 | 1,377 | 0,000 | 20 | 24 |
| Jacobs | 2019 | -0,074 | 0,819 | 20 | 18 |
| Jacobs_p | 2019 | -0,082 | 0,801 | 20 | 18 |
| Sorgdrager | 2019 | -0,104 | 0,671 | 32 | 35 |
| Sorgdrager_s | 2019 | -0,188 | 0,431 | 33 | 38 |
| Whiley | 2021 | 0,461 | 0,001 | 103 | 96 |
| Willette | 2021 | 0,019 | 0,907 | 112 | 58 |
| | | 0,021 | 0,835 | 580 | 503 |

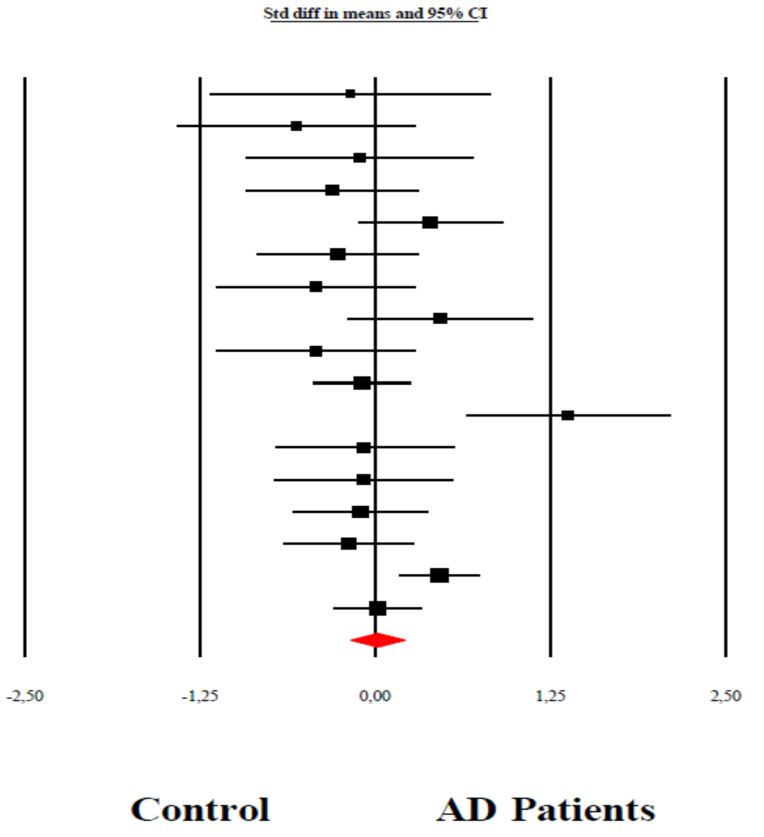

**Almulla et al, 2022**

**ESF, Figure 4:** The forest plot of 3-Hydroxykynurenine/kynurenine (KYN) ratio in patients with Alzheimer disease (AD) compared to healthy control reflecting KMO enzyme activity**.**

#### (KYN+3HK+QA+XA+PA)/(KA+AA)

| Study name | Year | Std diff in means | p-Value | Sample size AD-Patients | Sample size Control |
|---|---|---|---|---|---|
| Tohgi-b | 1992 | -2,084 | 0,000 | 14 | 10 |
| Tohgi-a | 1995 | -0,811 | 0,060 | 15 | 10 |
| Baran | 1999 | -0,426 | 0,305 | 11 | 13 |
| Widner | 2000 | 0,300 | 0,340 | 21 | 20 |
| Hartai | 2007 | 0,178 | 0,503 | 28 | 31 |
| Gulaj | 2010 | 0,419 | 0,158 | 34 | 18 |
| Greilberger | 2010 | 0,422 | 0,246 | 16 | 15 |
| Schwarz | 2013 | 0,346 | 0,293 | 20 | 19 |
| Wennstrom | 2014 | 0,024 | 0,939 | 19 | 20 |
| Rommer | 2015 | 0,422 | 0,246 | 16 | 15 |
| Giil | 2017 | -0,021 | 0,907 | 65 | 65 |
| Oxenkrug | 2017 | 1,308 | 0,001 | 20 | 24 |
| Jacobs | 2019 | -0,143 | 0,664 | 20 | 18 |
| Sorgdrager | 2019 | 0,152 | 0,539 | 32 | 35 |
| Velpen | 2019 | 0,017 | 0,943 | 40 | 34 |
| Jacobs_p | 2019 | 0,157 | 0,631 | 20 | 18 |
| Sorgdrager_s | 2019 | 0,058 | 0,807 | 33 | 38 |
| Sanchez | 2020 | -1,378 | 0,001 | 13 | 20 |
| Sanchez | 2020 | -0,073 | 0,843 | 13 | 20 |
| Whiley | 2021 | -0,029 | 0,840 | 103 | 96 |
| Willette | 2021 | -0,019 | 0,907 | 112 | 58 |
| | | 0,004 | 0,967 | 665 | 597 |

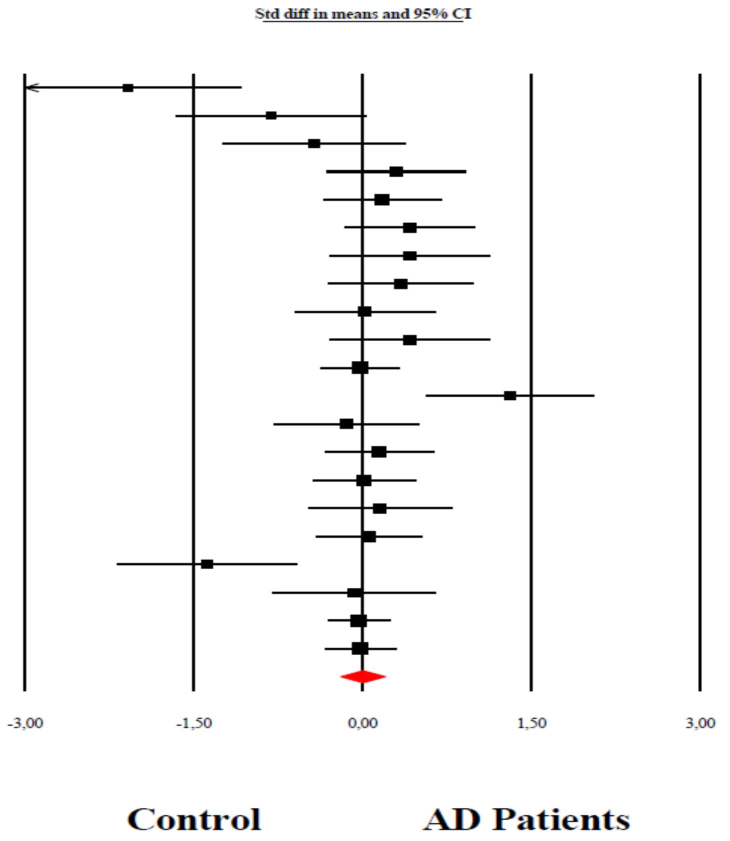

**Almulla et al, 2022**

**ESF, Figure 5:** The forest plot of kynurenine (KYN)+3Hydroxykynurenine (3HK)+Quinolinic acid(QA)+Xanthurenic acid (XA)+Picolinic acid (PA)/Kynurenine acid (KA)+Anthranilic acid(AA) ratio in patients with Alzheimer disease (AD) compared to healthy control reflecting neurotoxic over neuroprotective TRYCATs.

### KYN

| Study name | Year | Std diff in means | p-Value | Sample size: AD-Patients | Sample size: Control |
|---|---|---|---|---|---|
| Tohgi-b | 1992 | -1,909 | 0,000 | 14 | 10 |
| Tohgi-a | 1995 | -0,252 | 0,539 | 15 | 10 |
| Baran | 1999 | -0,289 | 0,483 | 11 | 13 |
| Widner | 2000 | 0,300 | 0,340 | 21 | 20 |
| Hartai | 2007 | -0,396 | 0,132 | 28 | 31 |
| Gulaj | 2010 | 0,596 | 0,045 | 34 | 18 |
| Greilberger | 2010 | 0,422 | 0,246 | 16 | 15 |
| Schwarz | 2013 | 0,307 | 0,340 | 20 | 19 |
| Rommer | 2015 | 0,422 | 0,246 | 16 | 15 |
| Giil | 2017 | -0,300 | 0,089 | 65 | 65 |
| Oxenkrug | 2017 | 0,100 | 0,740 | 20 | 24 |
| Jacobs | 2019 | 0,041 | 0,899 | 20 | 18 |
| Jacobs | 2019 | 0,283 | 0,387 | 20 | 18 |
| Sorgdrager | 2019 | -0,021 | 0,932 | 32 | 35 |
| Sorgdrager | 2019 | 0,039 | 0,870 | 33 | 38 |
| Whiley | 2021 | -0,461 | 0,001 | 103 | 96 |
| Willette | 2021 | -0,019 | 0,907 | 112 | 58 |
| | | -0,045 | 0,661 | 580 | 503 |

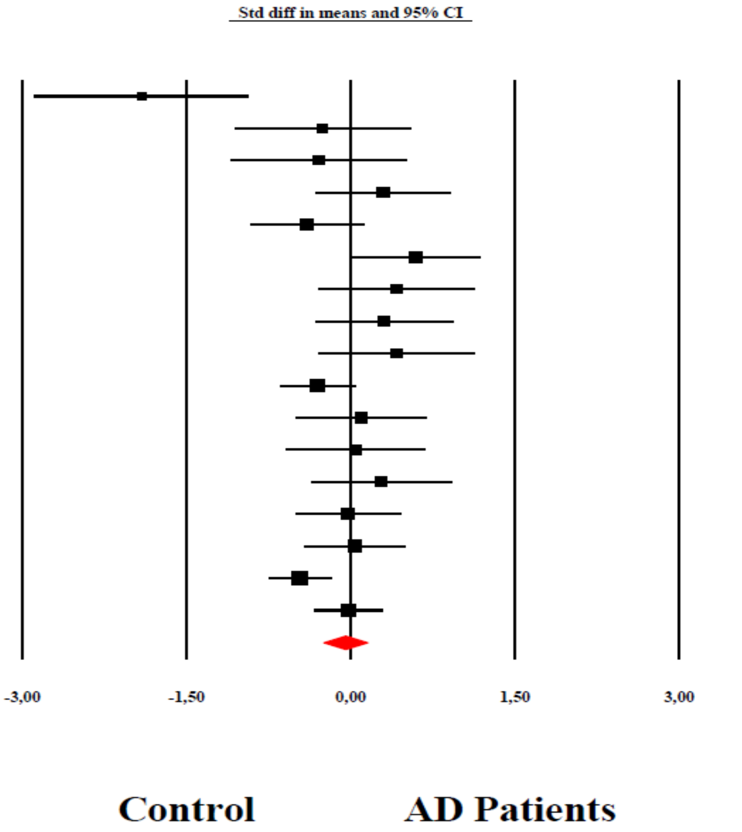

**Almulla et al, 2022**

**ESF, Figure 6:** The forest plot of kynurenine (KYN) in patients with Alzheimer disease (AD) compared to healthy control.

# KA

| Study name | Year | Subgroup within study | Std diff in means | p-Value | Sample size: AD-Patients | Sample size: Control |
|---|---|---|---|---|---|---|
| Baran | 1999 | CNS | 0,490 | 0,242 | 11 | 13 |
| Jacobs | 2019 | CNS | 0,972 | 0,005 | 20 | 18 |
| Sanchez | 2020 | CNS | 1,378 | 0,001 | 13 | 20 |
| Sorgdrager | 2019 | CNS | -0,693 | 0,006 | 32 | 35 |
| Velpen | 2019 | CNS | 0,283 | 0,227 | 40 | 34 |
| Wennstrom | 2014 | CNS | -0,024 | 0,939 | 19 | 20 |
| | | CNS | 0,363 | 0,227 | 135 | 140 |
| Giil | 2017 | Plasma | -0,364 | 0,040 | 65 | 65 |
| Gulaj | 2010 | Plasma | -0,991 | 0,001 | 34 | 18 |
| Hartai | 2007 | Plasma | -0,753 | 0,005 | 28 | 31 |
| Jacobs | 2019 | Plasma | -0,068 | 0,834 | 20 | 18 |
| Oxenkrug | 2017 | Plasma | -0,450 | 0,142 | 20 | 24 |
| Sanchez | 2020 | Plasma | 0,073 | 0,843 | 13 | 20 |
| | | Plasma | -0,448 | 0,002 | 180 | 176 |
| Schwarz | 2013 | Serum | -0,014 | 0,964 | 20 | 19 |
| Sorgdrager | 2019 | Serum | -0,227 | 0,341 | 33 | 38 |
| | | Serum | -0,151 | 0,429 | 53 | 57 |
| | | Overall | -0,249 | 0,021 | 368 | 373 |

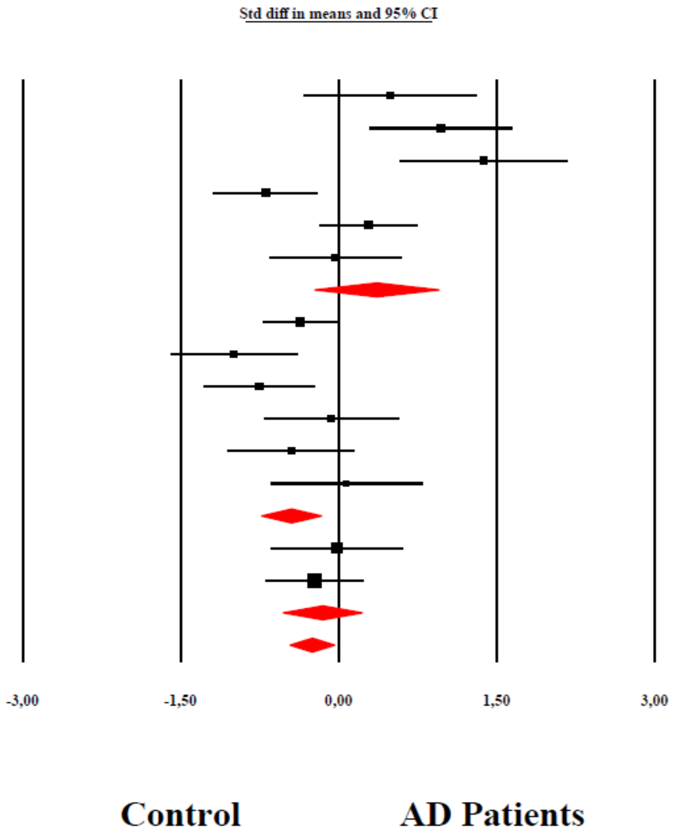

Almulla et al, 2022

**ESF, Figure 7:** The forest plot of kynurenic acid (KA) in patients with Alzheimer disease (AD) compared to healthy control.

## 3HK

| Study name | Year | Std diff in means | p-Value | Sample size: AD-Patients | Sample size: Control |
|---|---|---|---|---|---|
| Tohgi-b | 1992 | -2,259 | 0,000 | 14 | 10 |
| Tohgi-a | 1995 | -1,371 | 0,002 | 15 | 10 |
| Baran | 1999 | -0,501 | 0,229 | 11 | 13 |
| Gulaj | 2010 | 0,073 | 0,802 | 34 | 18 |
| Schwarz | 2013 | 1,245 | 0,000 | 20 | 19 |
| Giil | 2017 | -0,480 | 0,007 | 65 | 65 |
| Oxenkrug | 2017 | 2,855 | 0,000 | 20 | 24 |
| Jacobs | 2019 | -0,108 | 0,740 | 20 | 18 |
| Jacobs | 2019 | 0,118 | 0,716 | 20 | 18 |
| Sorgdrager | 2019 | -0,229 | 0,351 | 32 | 35 |
| Sorgdrager | 2019 | -0,337 | 0,160 | 33 | 38 |
| | | -0,072 | 0,801 | 284 | 268 |

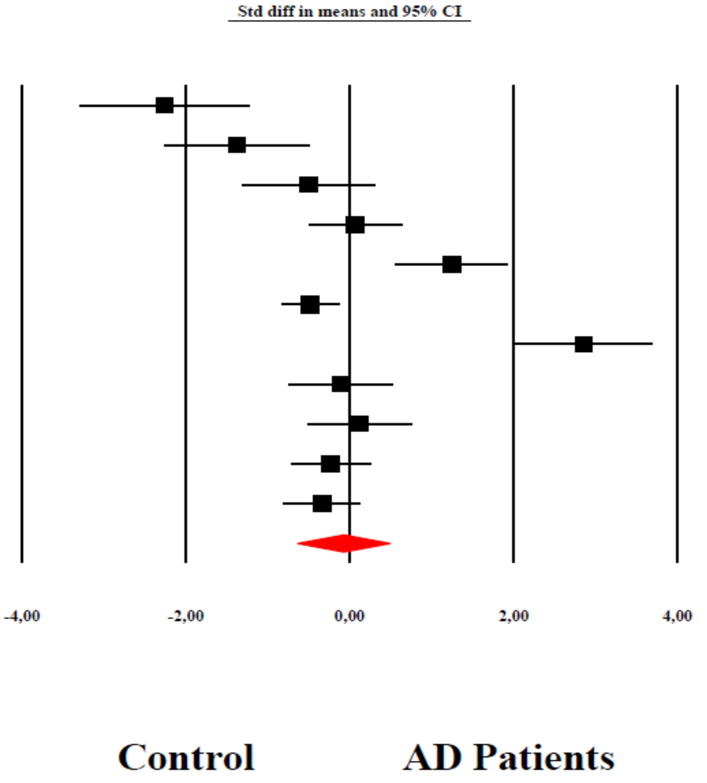

**Almulla et al, 2022**

**ESF, Figure 8:** The forest plot of 3-Hydroxykynurenine (3HK) in patients with Alzheimer disease (AD) compared to healthy control.

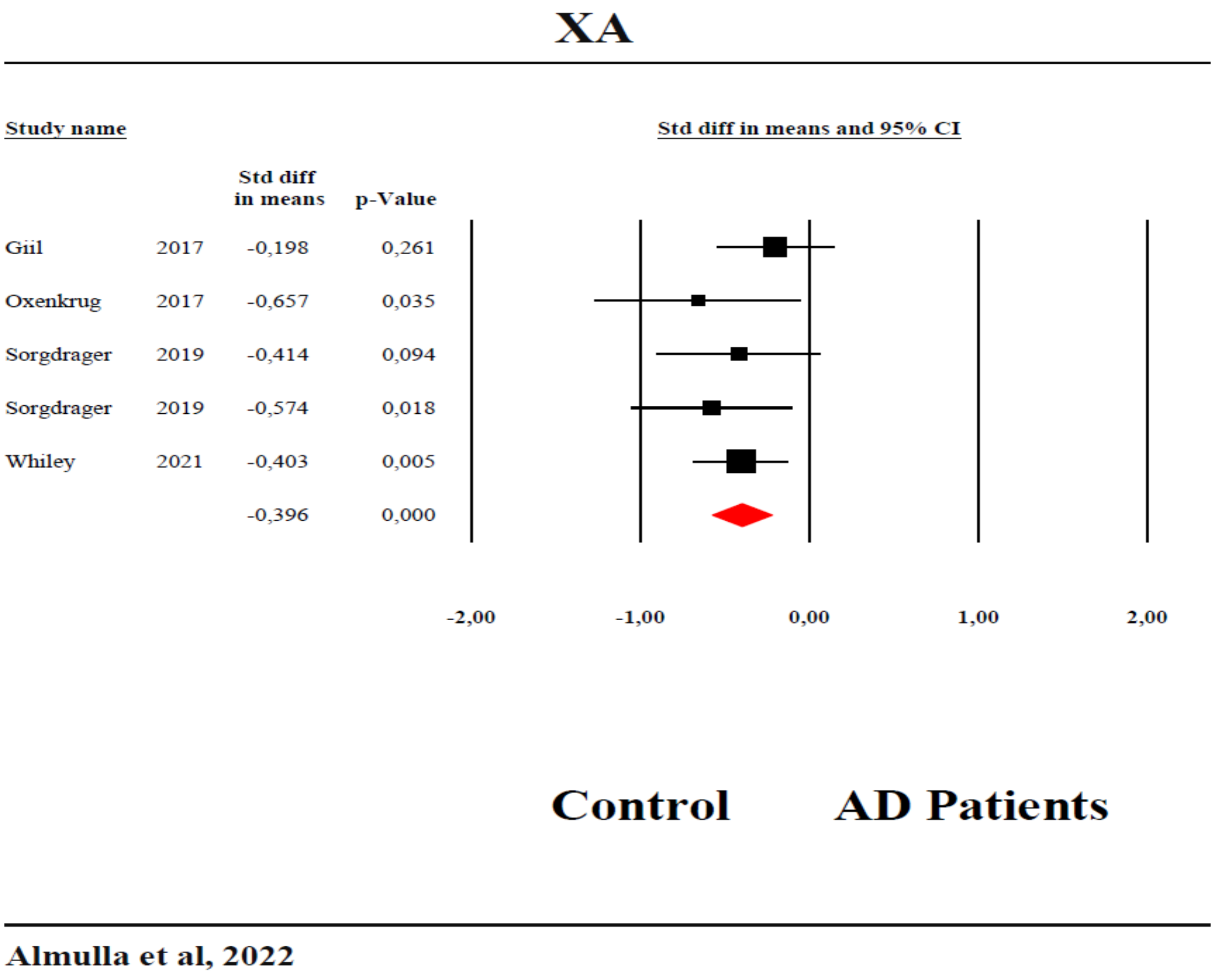

**ESF, Figure 9:** The forest plot of xanthurenic acid (XA) in patients with Alzheimer disease (AD) compared to healthy control.

**Abbreviations:**

AD: Alzheimer disease

TRP: Tryptophan

KYN: Kynurenine

KA: Kynurenic acid

3HK: 3-Hydroxykynurenine

AA: Anthranilic acid

3HA: 3-Hydroxyanthranilic acid

XA: Xanthurenic acid

QA: Quinolinic acid

PA: Picolinic acid

IDO: Indoleamine 2,3 dioxygenase

TDO: Tryptophan 2,3 dioxygenase

KAT: Kynurenine aminotransferase

KMO: Kynurenine 3-monooxygenase

KYNU: Kynureninase

$NAD^+$: Nicotinamide adenine dinucleotide

TRYCATs: Tryptophan Catabolites

TRYCAT pathway: Tryptophan catabolite pathway

KP: Kynurenine pathway

SMD: Standardized mean difference

CI: Confidence intervals

CSF: Cerebrospinal fluid

LC-MS: Liquid chromatography-mass spectrometry

LC-MS/MS: Liquid chromatography with two mass spectrometry

UHPLC-MS: Ultra-high-performance liquid-chromatography- mass spectrometry

NMDA: N-methyl-D-aspartate

Aβ: amyloid-beta

IL-6: Interleukin-6

TNF-α: Tumor necrosis factor

IL-1ra: IL-1 receptor antagonist

IL-10: Interleukin-6

IL-1β: Interleukin-1beta

IFN-γ:  Interferon-gamma

O&NS: Oxidative and nitrosative stress

ROS: Reactive oxygen species

LPS: Lipopolysaccharides

VGLUT: Reducing vesicular glutamate transport

PRISMA: Preferred Reporting Items for Systematic Reviews and Meta-Analyses

MOOSE: Reviews and Interventions and Meta-Analyses of Observational Studies in Epidemiology

ESF: Electronic supplementary file

SD: Standard deviation

SEM: Standard error

ICS: Immune confounder scales

CNS: Central nervous system

NADPH: Nicotinamide adenine dinucleotide phosphate
